# Child public health interventions for conflict-affected populations: A systematic review

**DOI:** 10.1101/2025.06.18.25329756

**Authors:** Ayesha Kadir, Sneha Krishnan, Catherine McGowan, Daniel Martinez Garcia

## Abstract

**Background:** Armed conflict causes pervasive harm to children, and humanitarian responses to support them face significant challenges. This review aims to summarise the evidence on the effectiveness of interventions to treat, protect, and promote child public health in conflict-affected populations.

**Methods:** A systematic review was performed, with searches of major databases and the grey literature from 1 January 2012 – 20 February 2025. Included studies provided data on child or caregiver outcomes associated with interventions to support children affected by armed conflict. Studies on nutrition and perinatal interventions were excluded. Data were extracted on the setting, population, intervention design, study type, and findings.

**Results:** The searches yielded 3,601 records. 51 intervention studies met inclusion criteria, 39% of which were trials. Studies were mainly from Africa (51%), the Middle East (25%) and Asia (18%). The majority of studies focused on mental health and psychosocial support (MHPSS) (N=29, 57%). MHPSS, child protection, and/or parenting interventions were the focus of trials as well as intersectoral interventions. Somatic child health interventions (N=19, 37%) focused on immunisation, adolescent sexual and reproductive health, toxic stress, and telemedicine services. Five studies measured development outcomes and one intervention targeted children with disabilities. Over half of the studies were carried out amongst displaced populations. Intervention design varied widely within and between sectors. Studies showed promising results, particularly for non-specialist MHPSS interventions. Only 20% of studies assessed intervention safety.

**Conclusion:** The evidence for child public health interventions to support conflict-affected populations is increasing, with increased numbers of studies over time, and improved study design, execution, and reporting. However, the evidence remains poor, limited to a few topic areas and with continued geographical disparities. There is a lack of studies from Central and South America, the Caribbean, North Africa, West Africa, Southeast Asia, and the Pacific. There are notable gaps in evidence on the safety of interventions, their medium- and long-term impacts, sustainability, and interventions for child development and children living with disabilities.

## Introduction

Armed conflict is known to cause pervasive harm to children, including physical, psychological, and social injuries that can impact their health, psychomotor development, and life trajectories. The scale of the impact of armed conflict on children is large: 468 million children (or 1 in 6) live within 50 km of an active conflict zone, and more than 2/3 live in a country that is affected by armed conflict [1]. Conflict can impact children directly through combat activities, and indirectly through a range of pathways, including but not limited to destruction of infrastructure, forced displacement, lack ability to meet basic needs, and changes in social arrangements, all of which place children at heightened risk for experiencing violence, deprivation, and exploitation [2]. Conflicts also affect children who live far from where the fighting occurs. For example, family members may participate in or be directly affected by armed conflict; children living in other countries may witness news, social media imagery, and discourse about conflicts, attacks and violence; and they may experience economic and social changes that are related to increasingly internationalised conflicts [2].

The past two decades have seen increased interest in the impacts of conflict on health, and recent reviews have described the effects of armed conflict on child health [2–4]. These reviews suggest that conflict affects children through nuanced and context-specific ways, however the nuances remain poorly understood, thus limiting our understanding of the kinds of interventions that are appropriate, safe, and effective to mitigate harm and improve child health. There is marked geographic disparity in which populations of children are studied [2], and routine data on child public health in conflict settings are limited [5]. As such, we have sought to improve our understanding of the effectiveness and safety of recent interventions to support child health in conflict settings.

We report below the findings from a systematic review of intervention studies aimed at mitigating harm and improving child health. We report against items in the updated PRISMA reporting guidelines [6]. The review protocol is registered with PROSPERO (CRD42022356007).

## Methods

The exposure of interest was direct or indirect experience of armed conflict, including displacement due to conflict, living in an area where active combat is occurring or has occurred within the previous generation, or having a first degree relative who has been directly affected by armed conflict. We relied on author determinations of indirect exposure. Studies presenting data on child public health outcomes relating to conflict or interventions to support children impacted by conflict were included in the original review.

Database searches were conducted in MEDLINE, EMBASE and Global Health (all via Ovid) on 30 August 2022 using variations of terms for children, armed conflict, and humanitarian or crisis. Searches were limited to sources published since 1 January 2012. No language restrictions were applied. Duplicates were removed in EndNote [7] and the remaining records were uploaded to Covidence [8] for title/abstract screening and full text review.

Grey literature searches were conducted in DuckDuckGo, ReliefWeb, and in the websites of Médecins Sans Frontières, Save the Children International, International Rescue Committee and UNICEF on 27-28 September 2022 using the search terms “child” and “armed conflict”. The first 100 records retrieved from each search were screened on title/abstract, and subsequently on full text. Additional records were identified from review of citation lists during full text screening. Screening and full text review were completed independently by two reviewers. Discrepancies in screening outcomes were resolved through discussion. A third author resolved any persisting discrepancies.

We carefully considered carrying out a risk of bias assessment but ultimately opted not to. Conflict settings place extreme limits on public health research. Security risks, barriers in access to affected populations, and movements of populations typically compromise study rigour and introduce bias. However, omitting or otherwise reducing the importance of studies in conflict settings on the basis of study quality would, paradoxically, introduce bias towards less severe conflict settings and compromise external validity. Moreover, subjecting studies to a risk of bias assessment would almost certainly have required us to redefine the scope of our review. Finally, our review presents a typology of interventions, and intervention designs, aimed at improving the health and wellbeing of conflict-affected children; the identified studies did not enable meta-analysis or other quantitative synthesis. The application of gold standard criteria for studies in conflict-affected populations limits the availability of evidence and undermines the value of evidence from the hardest to reach populations [5, 9]. Lack of data renders these populations invisible, limits the ability of humanitarian actors to deliver safe and effective interventions, and hampers progress and improvement in research among conflict-affected populations. Taken together, the justification for carrying out a risk of bias assessment or a certainty assessment was weak when compared to the benefits of not doing so.

Studies were included if they were undertaken in humanitarian settings caused by armed conflict and reported substantively on interventions targeting children <18 years of age or their caregivers. Studies were included if they reported on any of the following outcomes: morbidity, disability, child development, child mental health, caregiver mental health, changes in social behaviours, schooling, and/or mortality. Papers that provided non-disaggregated data that included children <18 years as well as young people up to <24 years were included. In addition to original research articles, agency technical reports were included if a clear description of the study methods was provided. No restrictions were made for language or geographic location. Military studies were excluded, as these studies report on a special population that sought care from a military health facility, and are thus unlikely to represent the general population of children affected by conflict [2]. Studies that only provided data on nutrition, perinatal, or neonatal interventions and outcomes were excluded, due to recent reviews on these subjects [10–12].

An extraction form was created in Microsoft Excel [13] and included information about: setting, intervention design, type of conflict, location of study, study design, outcomes, and limitations. Two authors supported data extraction, one of whom reviewed the entire dataset for internal consistency in data extraction. A template data extraction form and analytic codes are available in the supplementary materials.

One author updated the MEDLINE and EMBASE searches on 20 February 2025, screened the titles using the original criteria but limited to intervention studies only, and extracted the findings for studies meeting inclusion criteria. A second author reviewed the dataset of the search update for internal consistency, including review of the full text papers.

The extracted data on intervention studies from the original review and the search update were compiled in Microsoft Excel, coded and analysed for trends in study design, geographic location, thematic focus, target group age and gender, displacement status, and whether the study mentioned exploration of any harm caused to children from the intervention. As the included studies varied markedly in focus, design, and reporting of outcomes, we were unable to carry out a meta-analysis.

## Results

The database searches retrieved 3094 records (Figure 1). After removing duplicates, 2108 records were screened, of which 235 were sought for retrieval. One full text paper was not available. 234 papers were reviewed in full text, of which 109 were excluded. The grey literature searches retrieved 501 records. Six additional papers were identified by hand searches of citations. After removing duplicates, 460 records were assessed for eligibility from the grey literature and citation searches. Our final sample included 51 intervention studies. The included studies and their main findings are presented in Table 1.

**Figure 1.** PRISMA flow diagram.

**Table 1.**
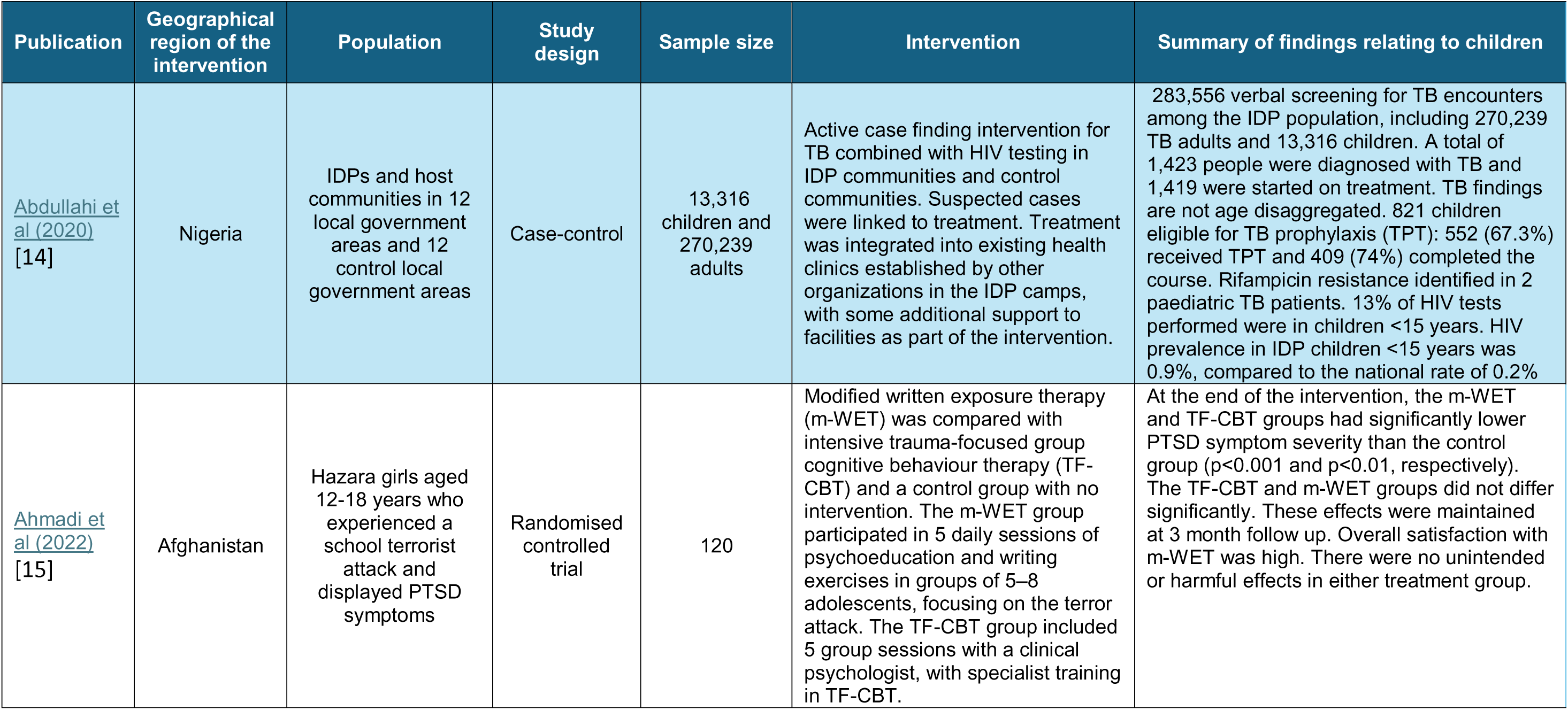
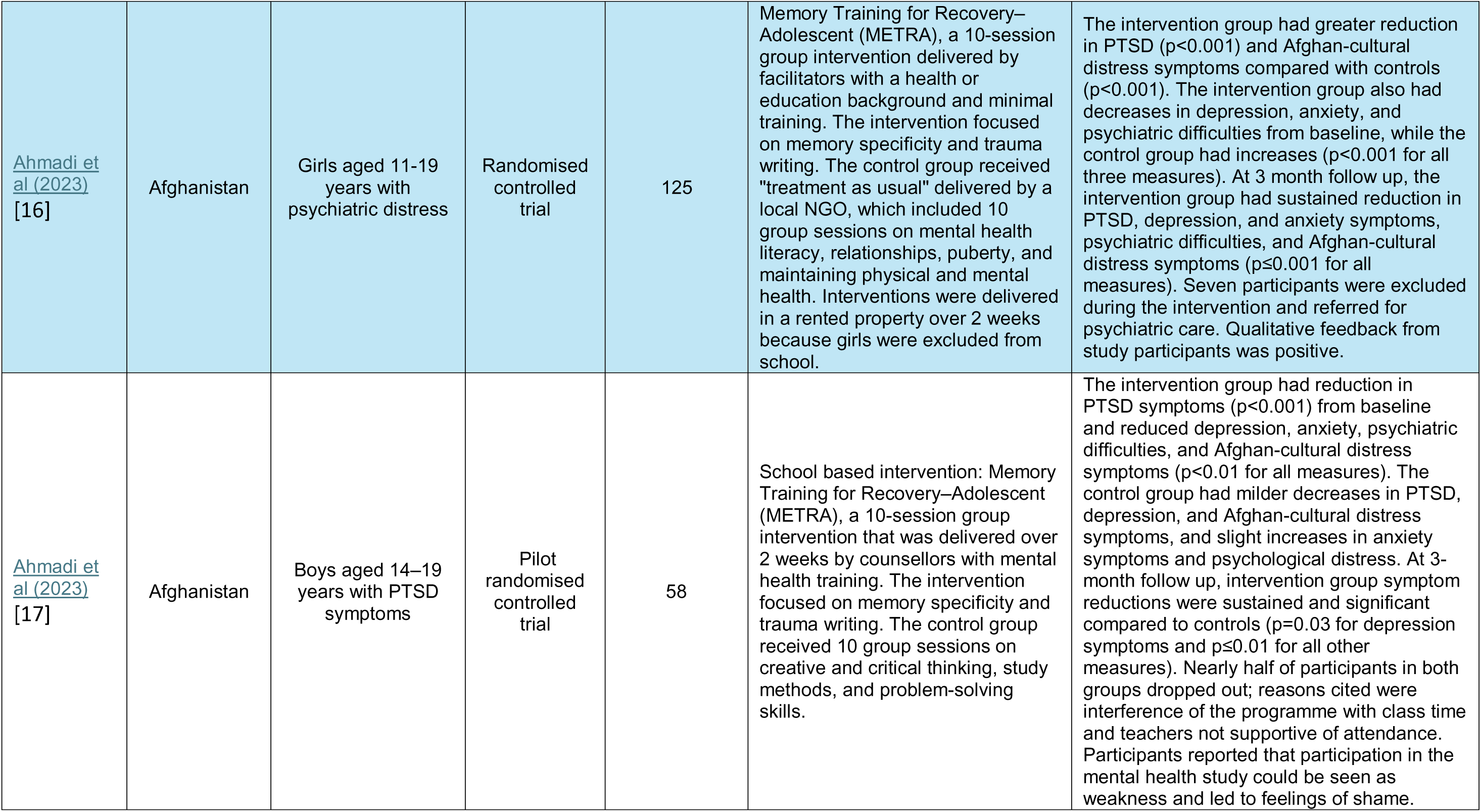
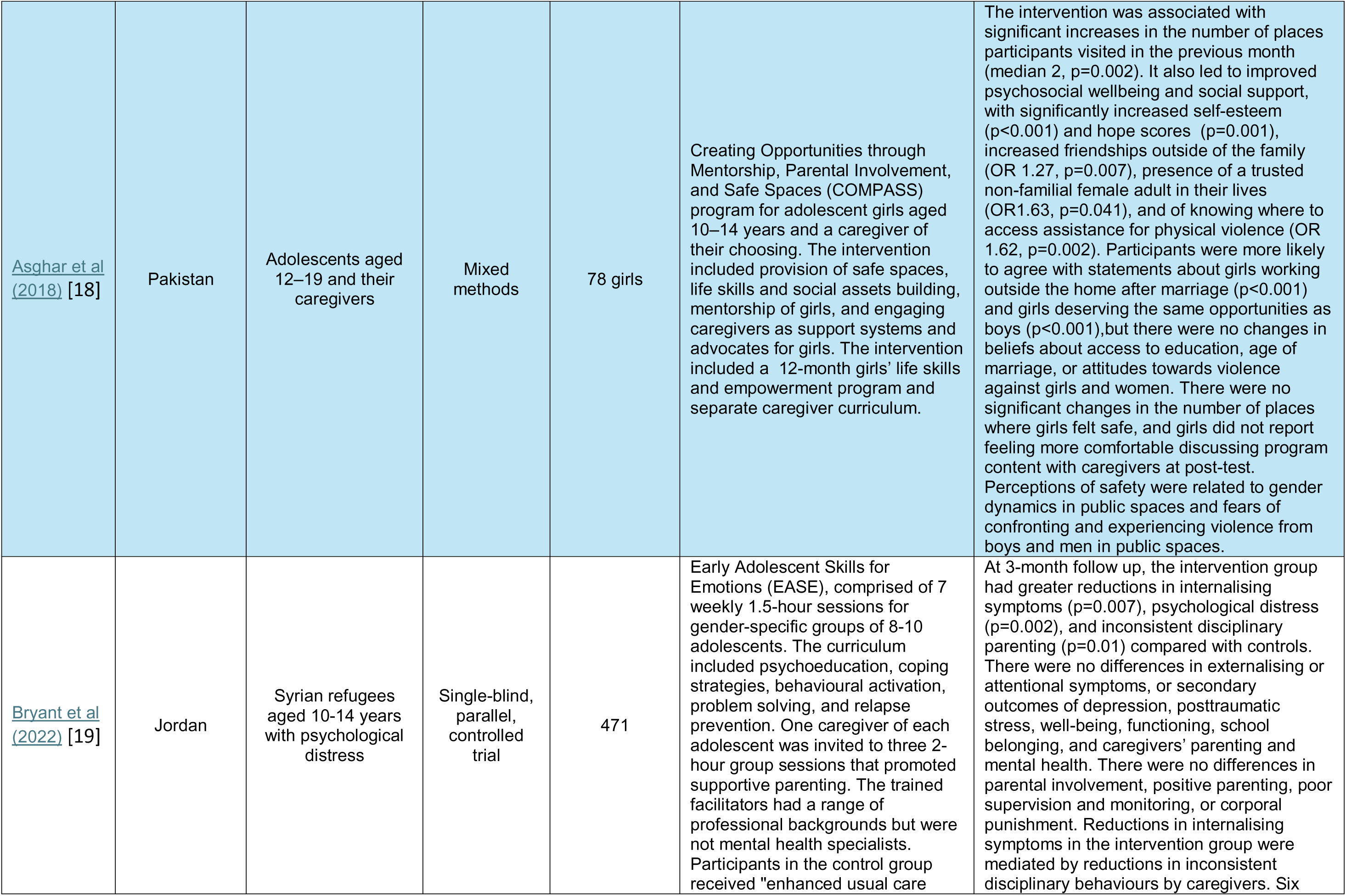
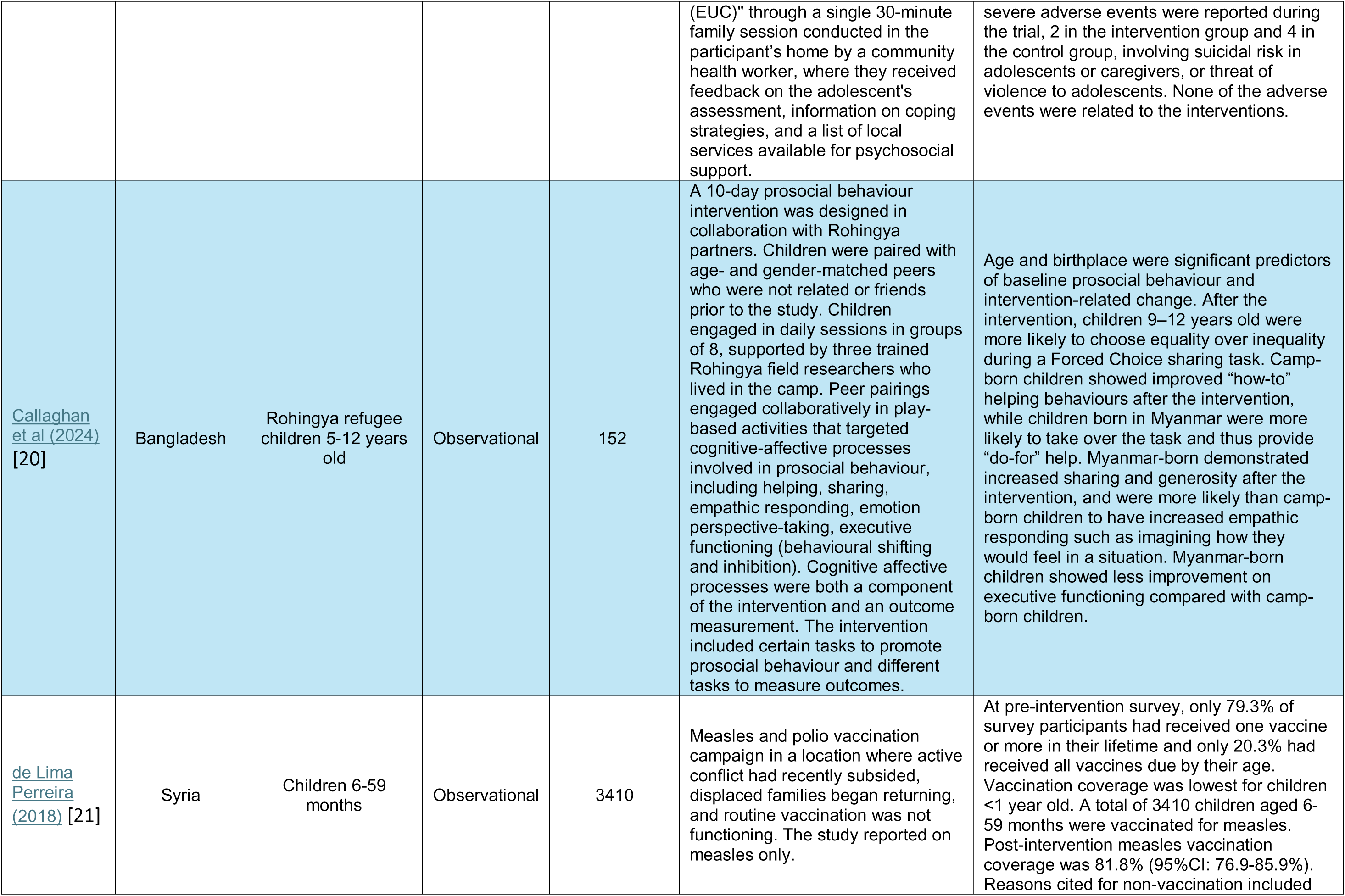
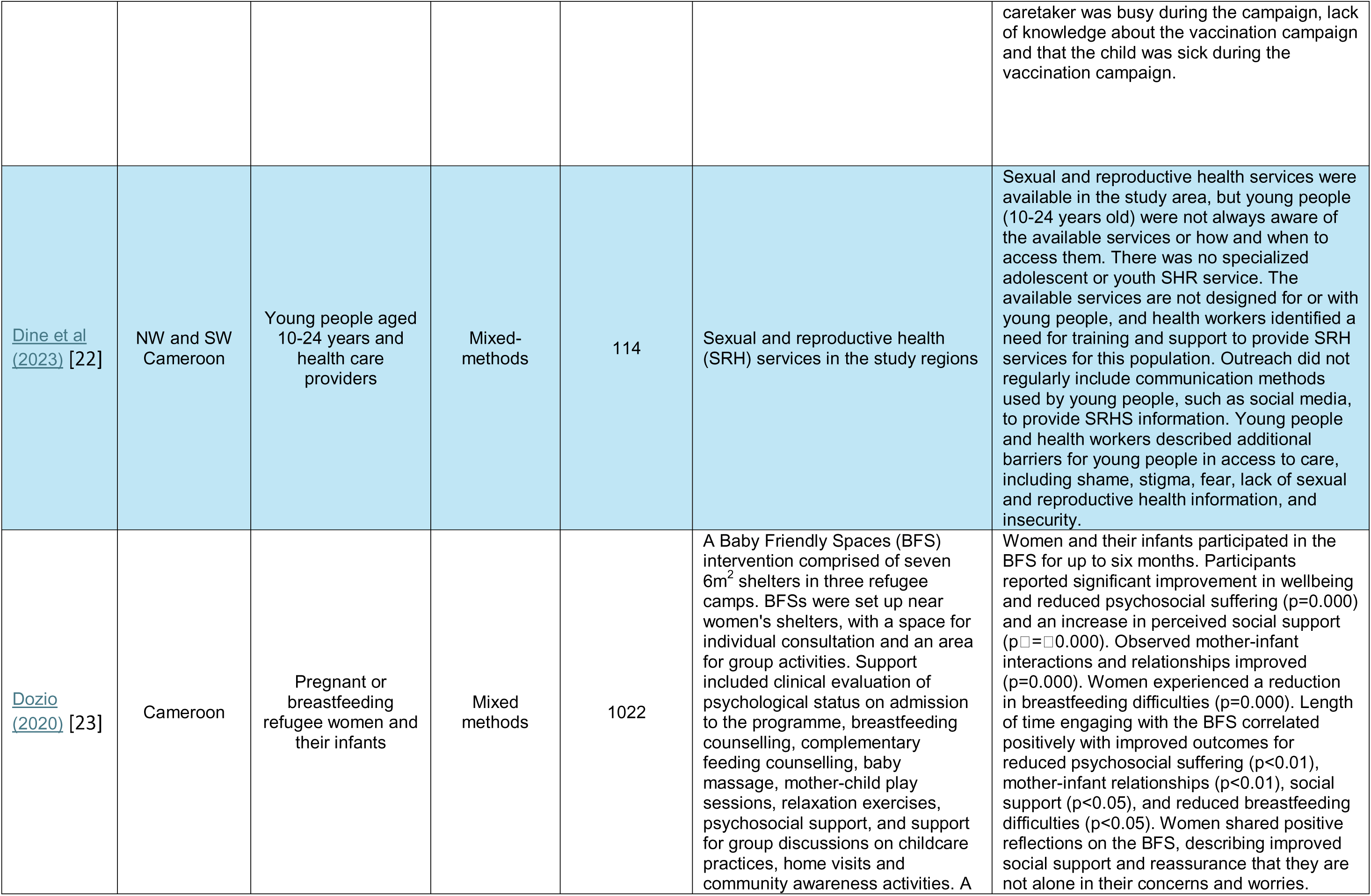
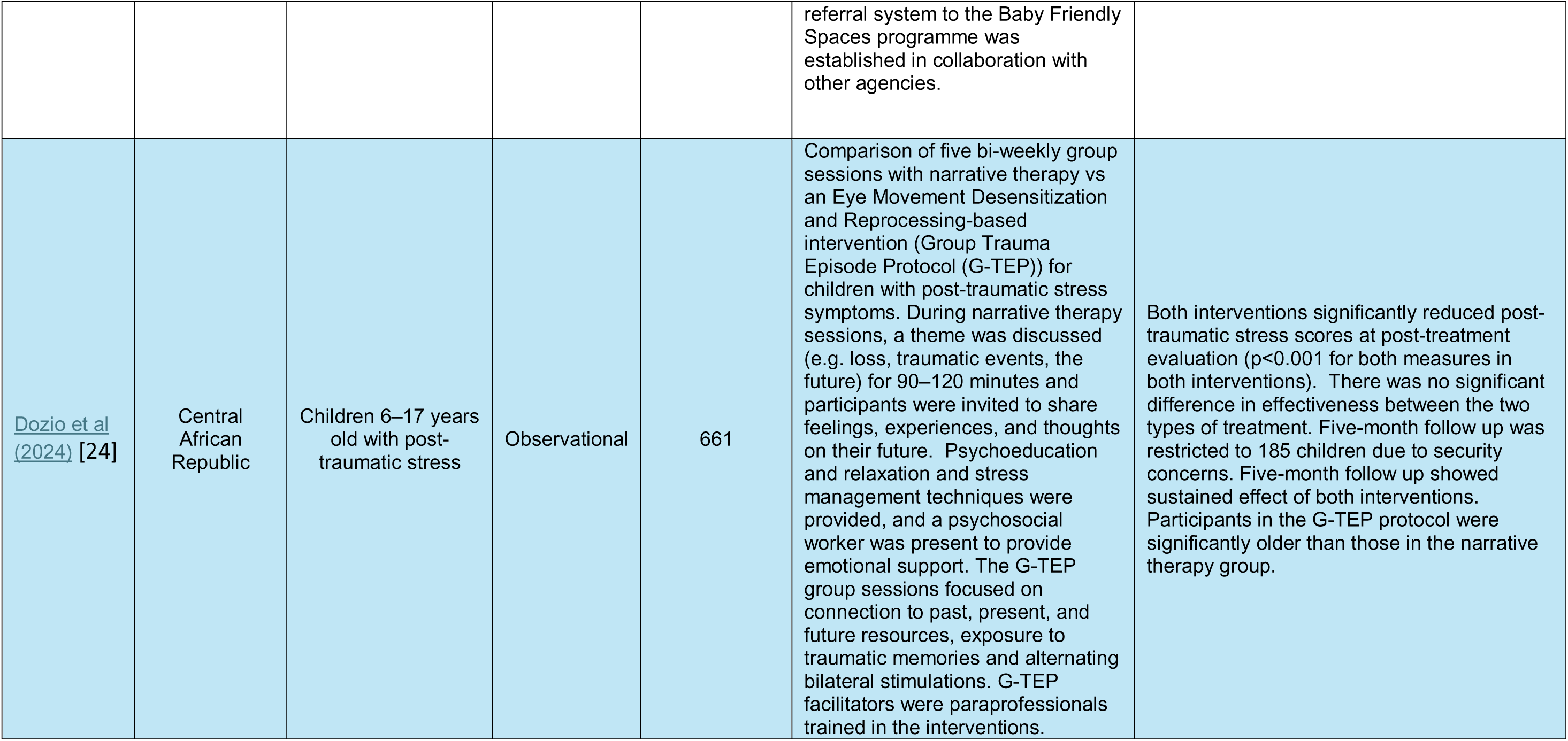
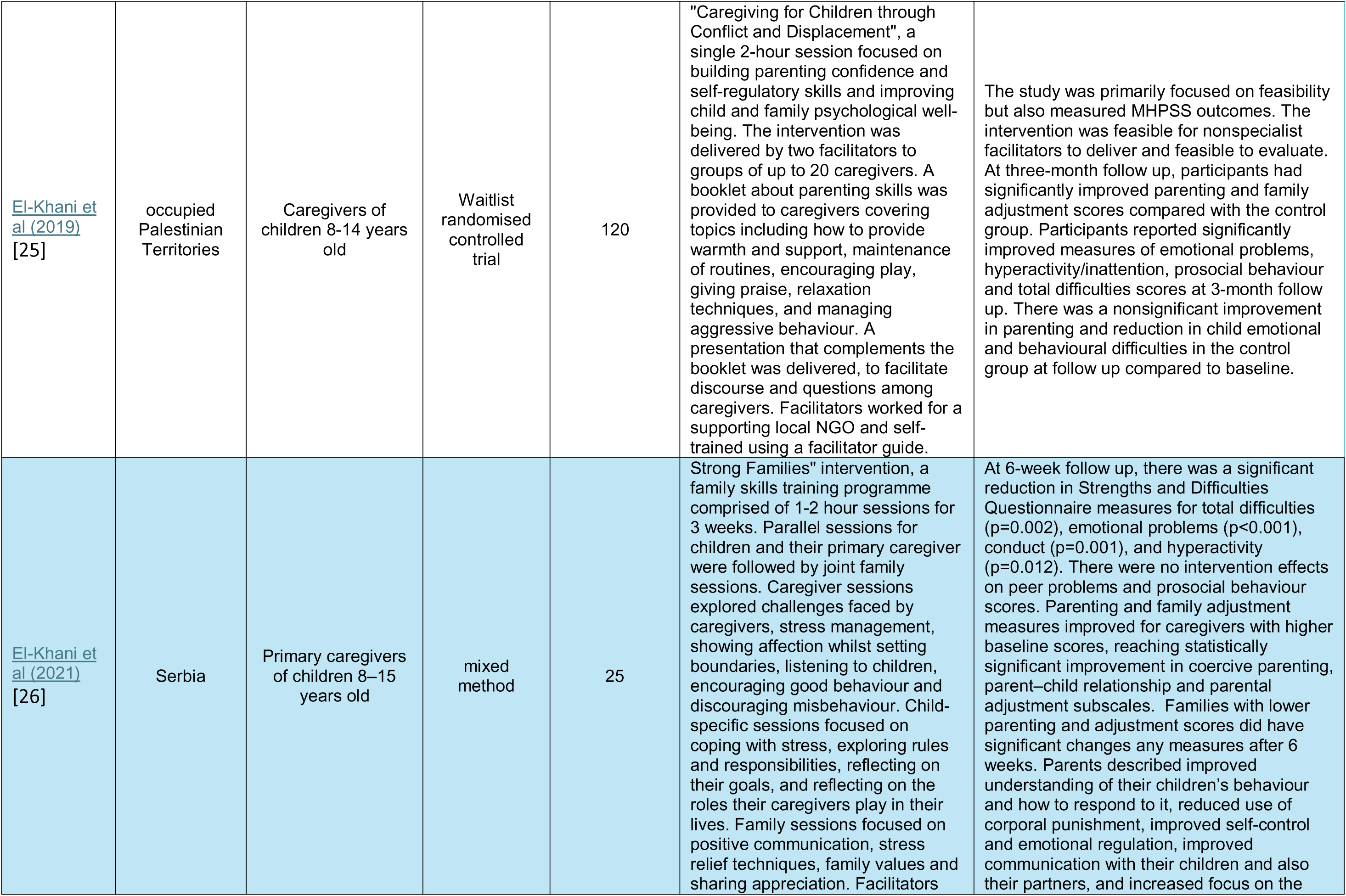
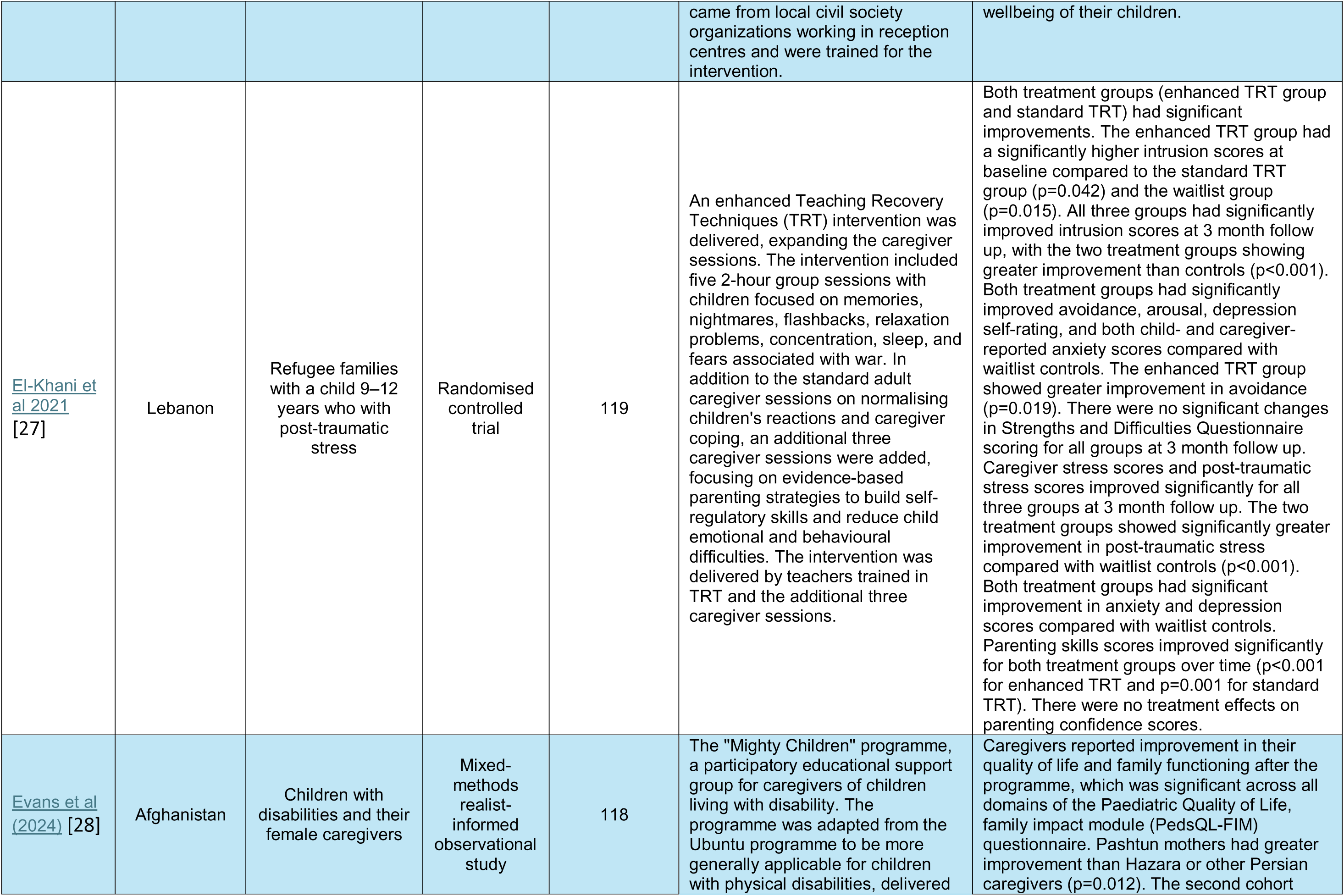
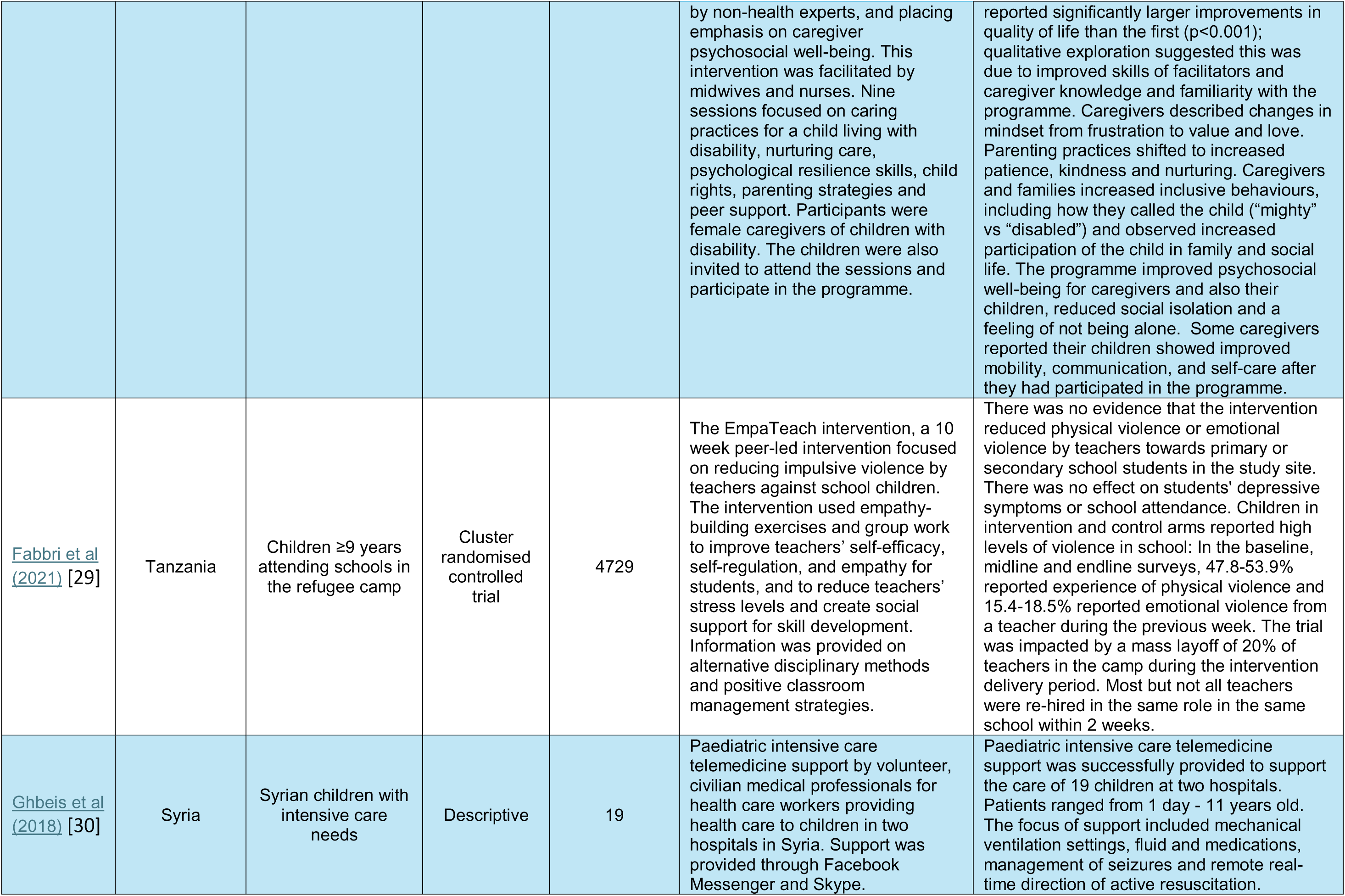
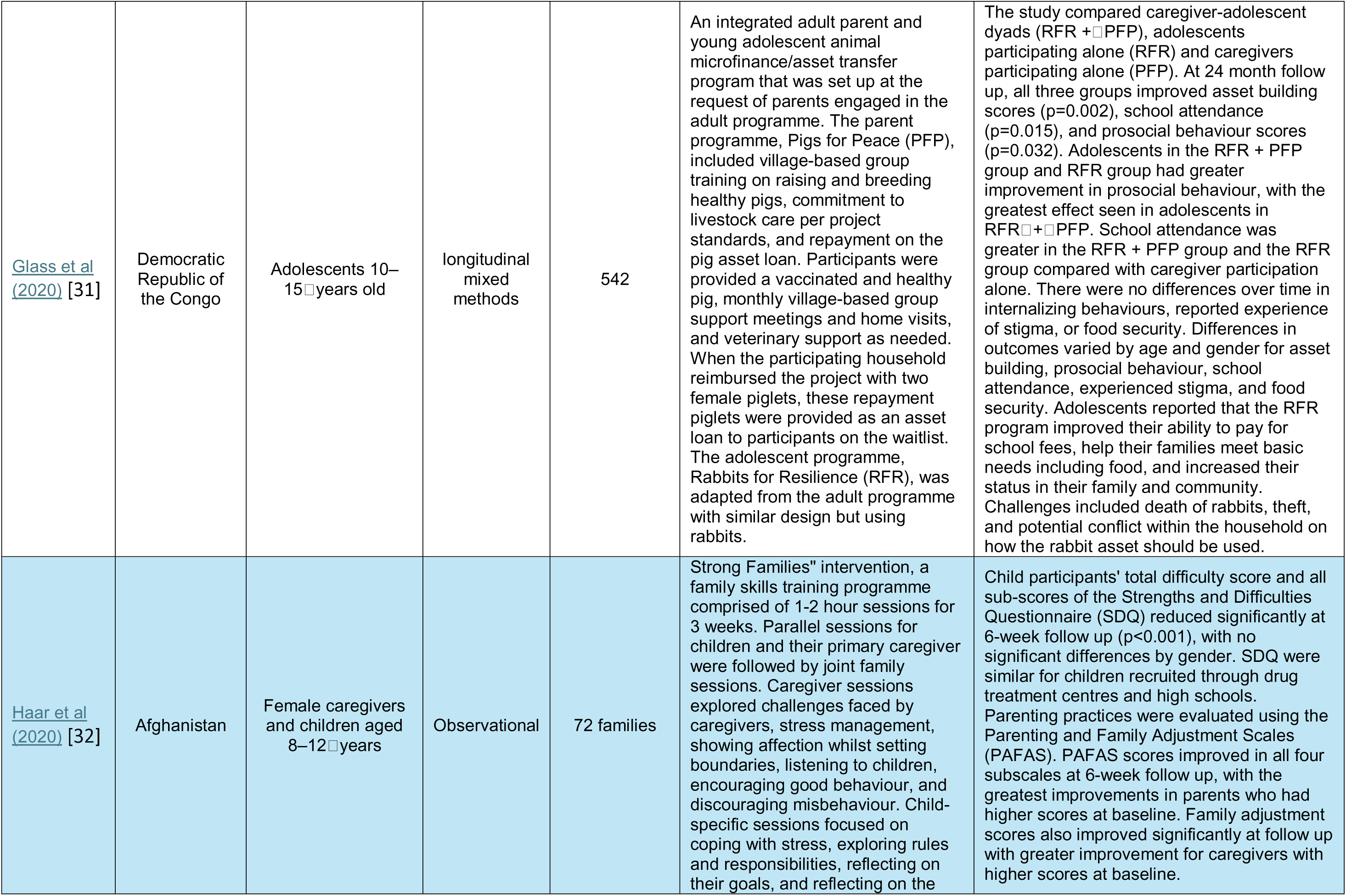
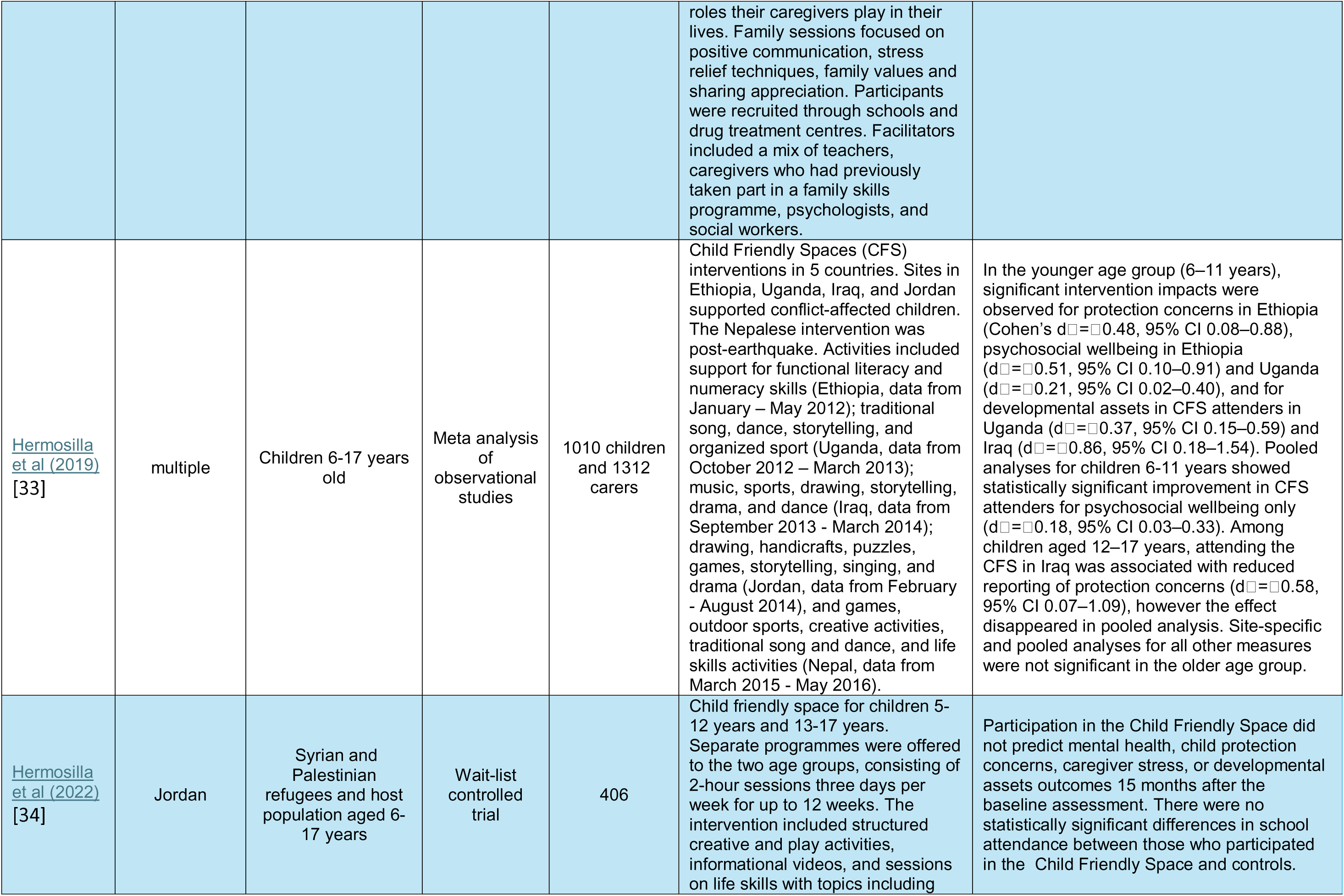
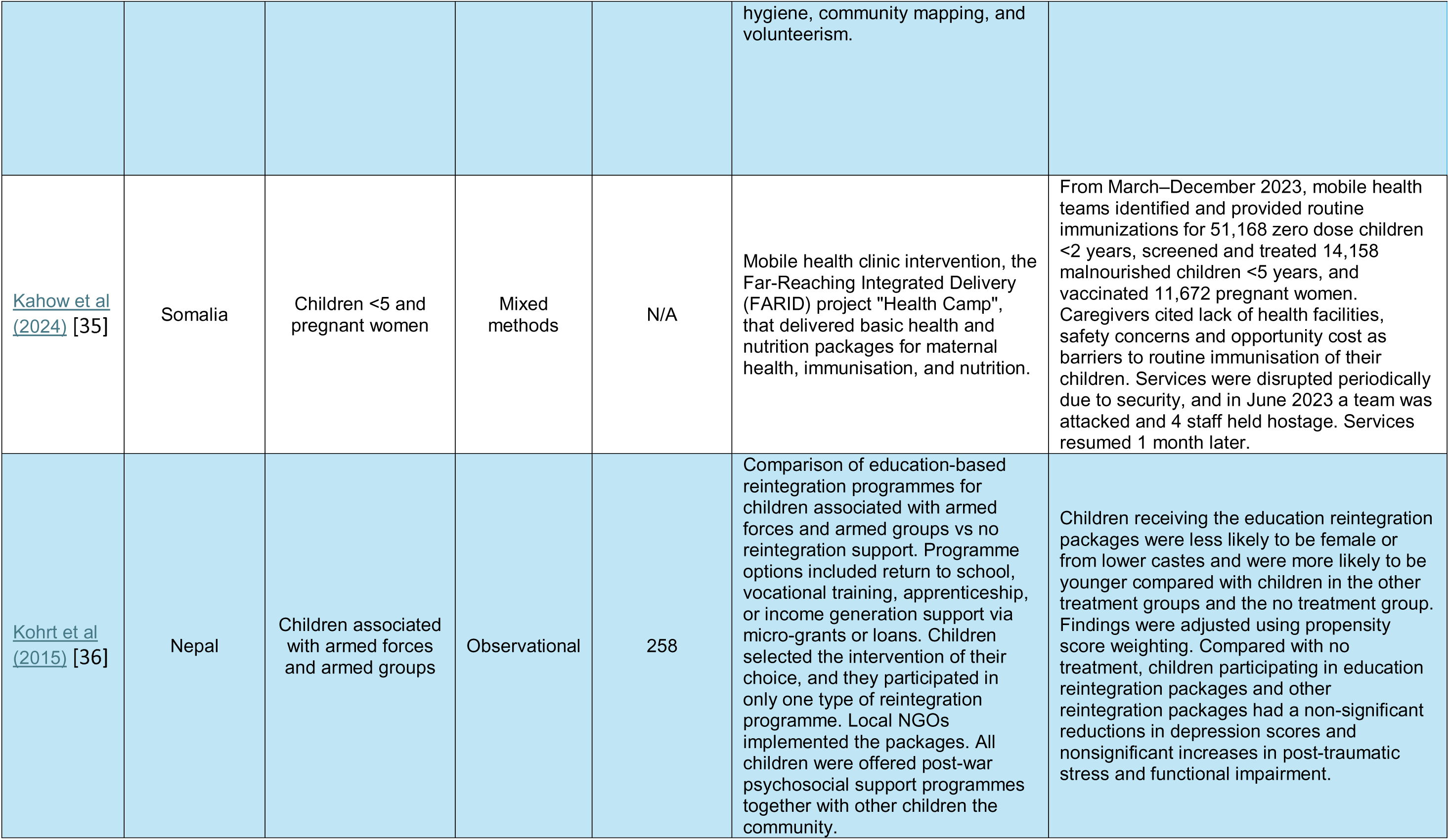
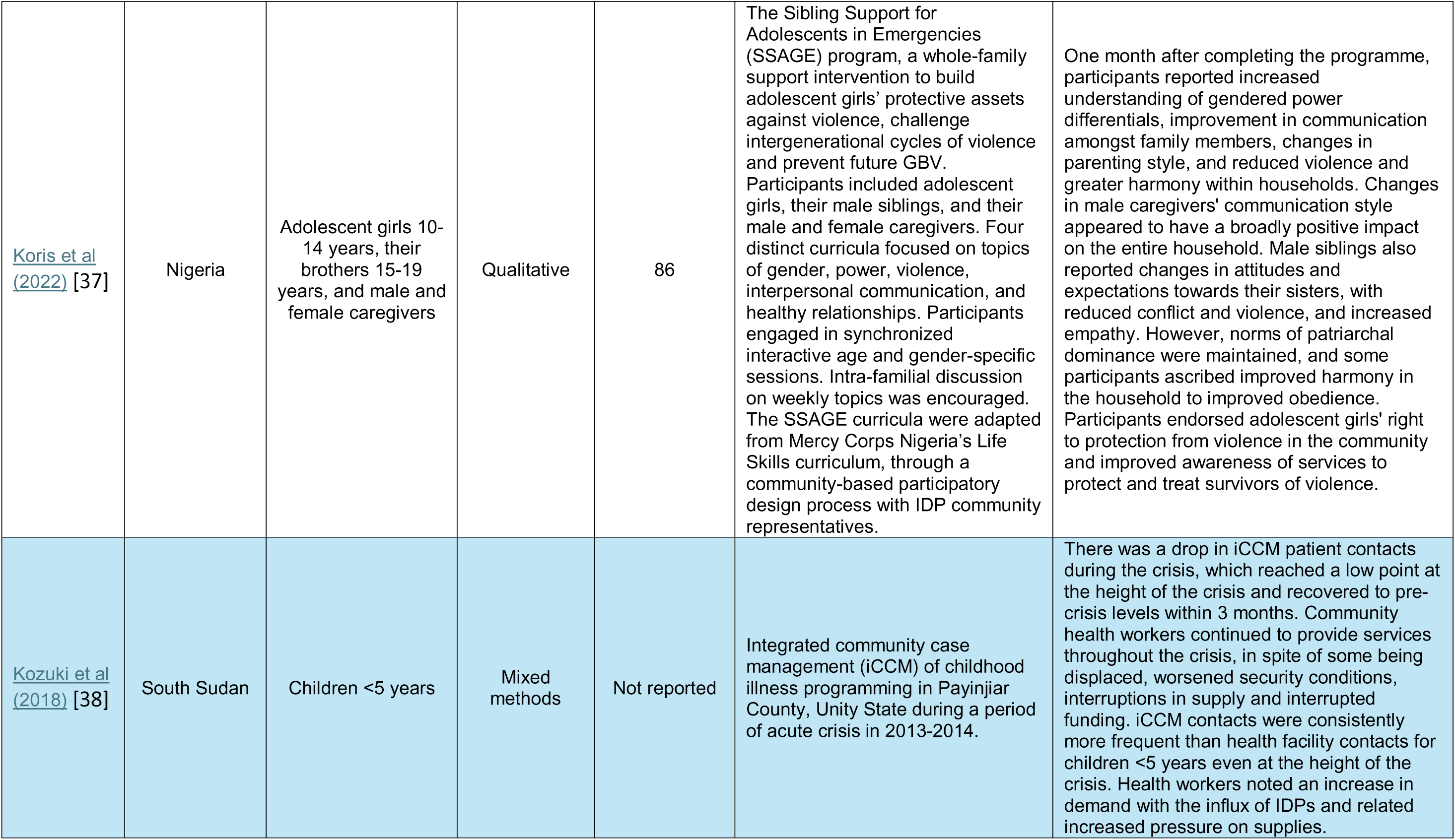
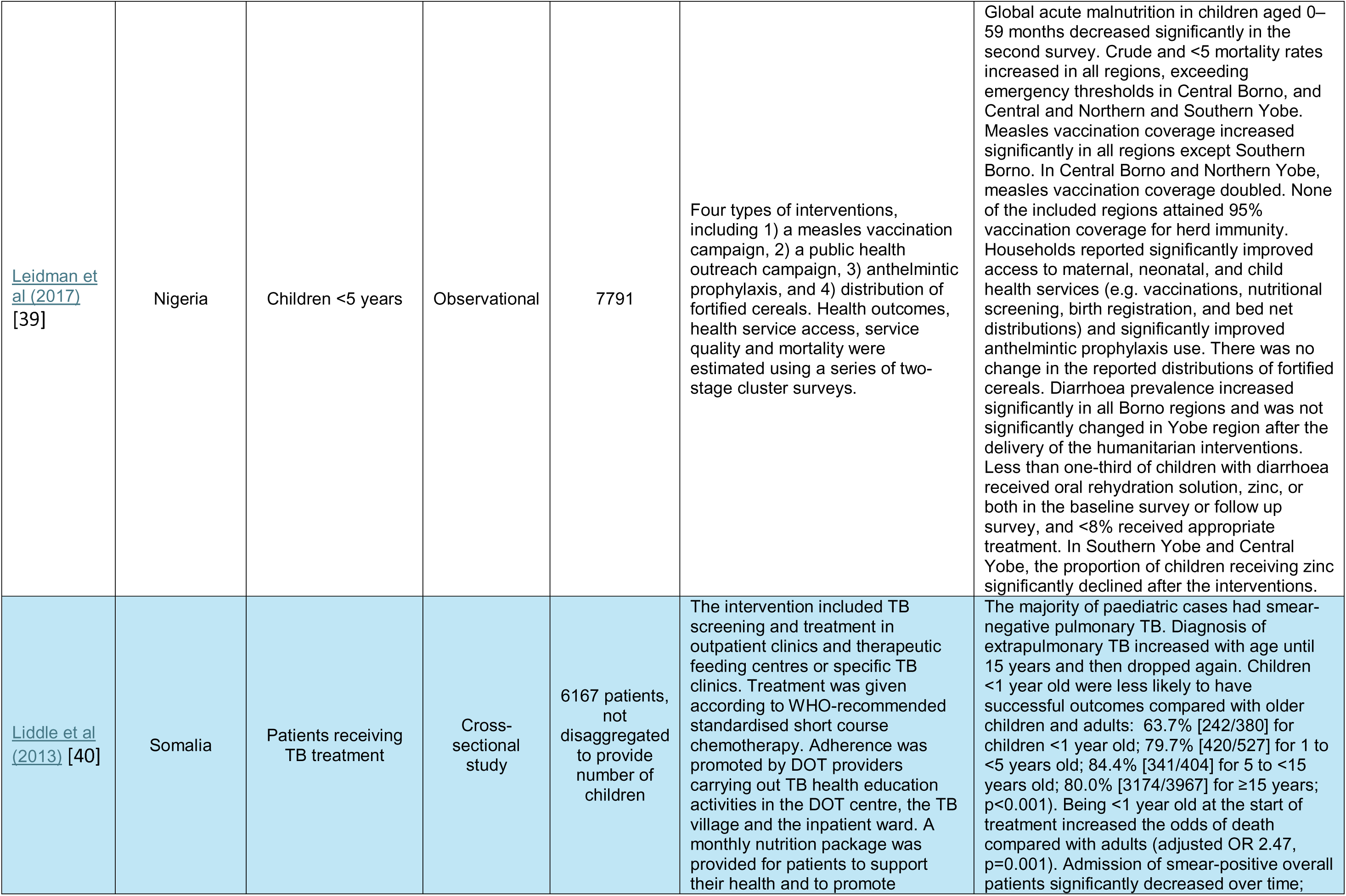
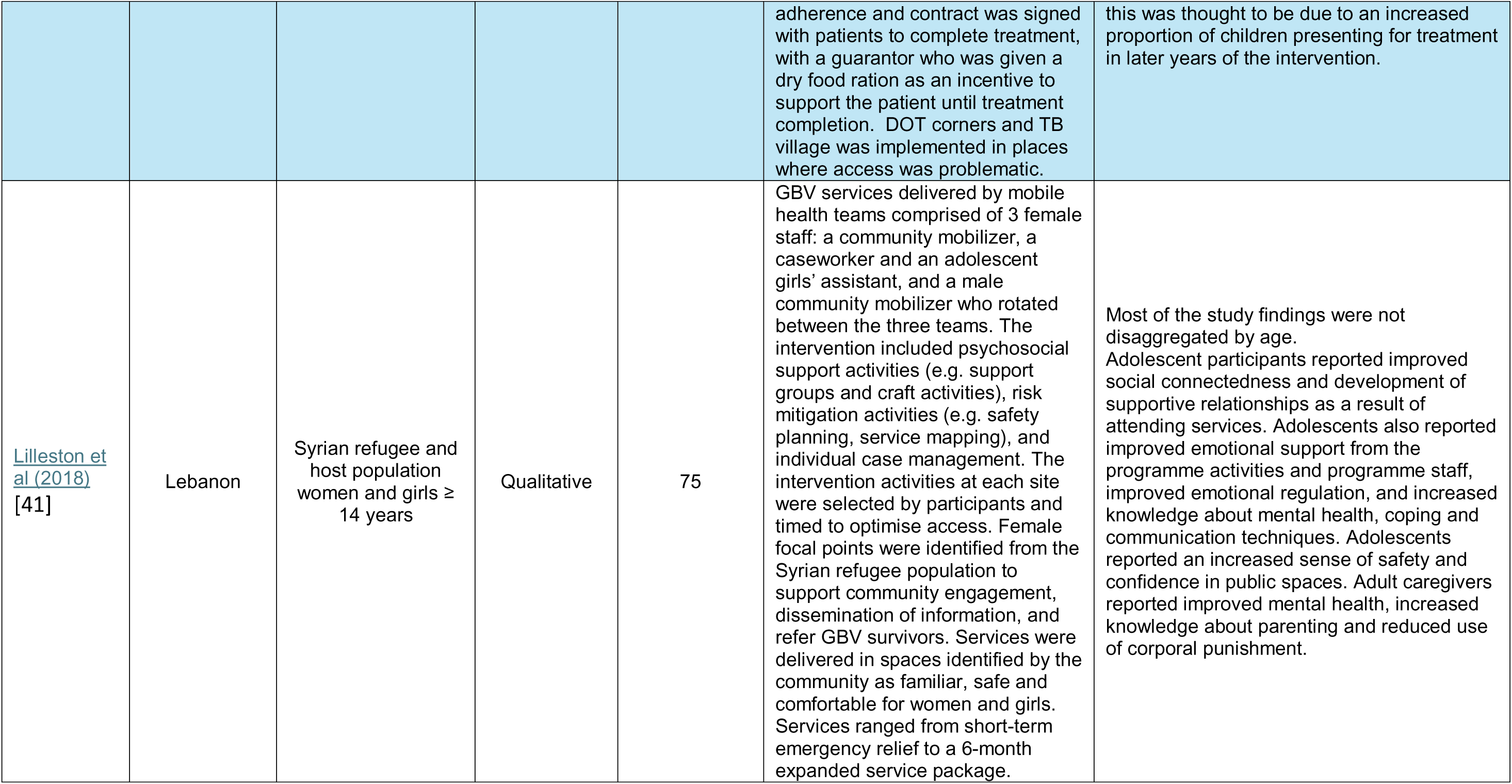
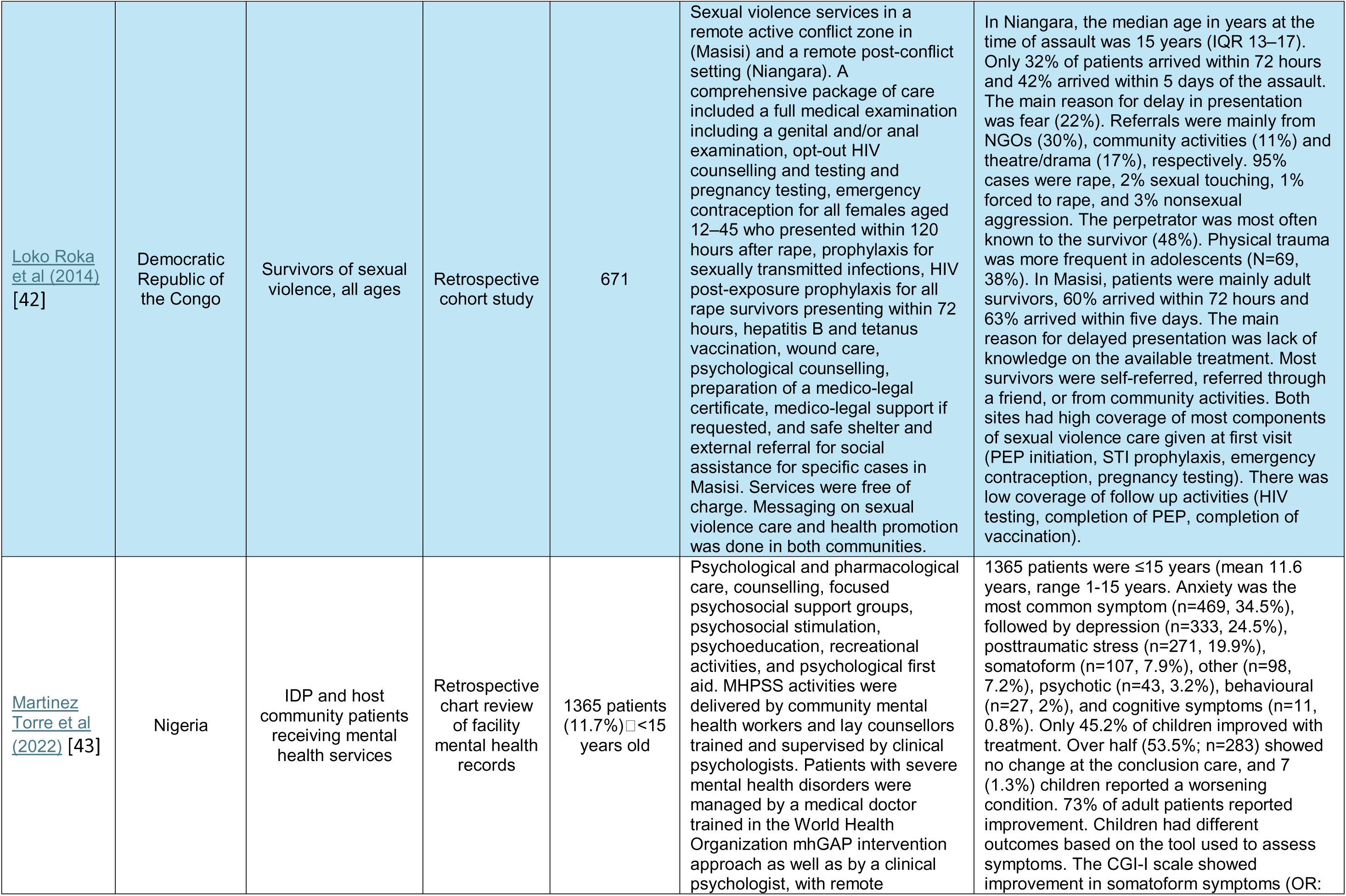
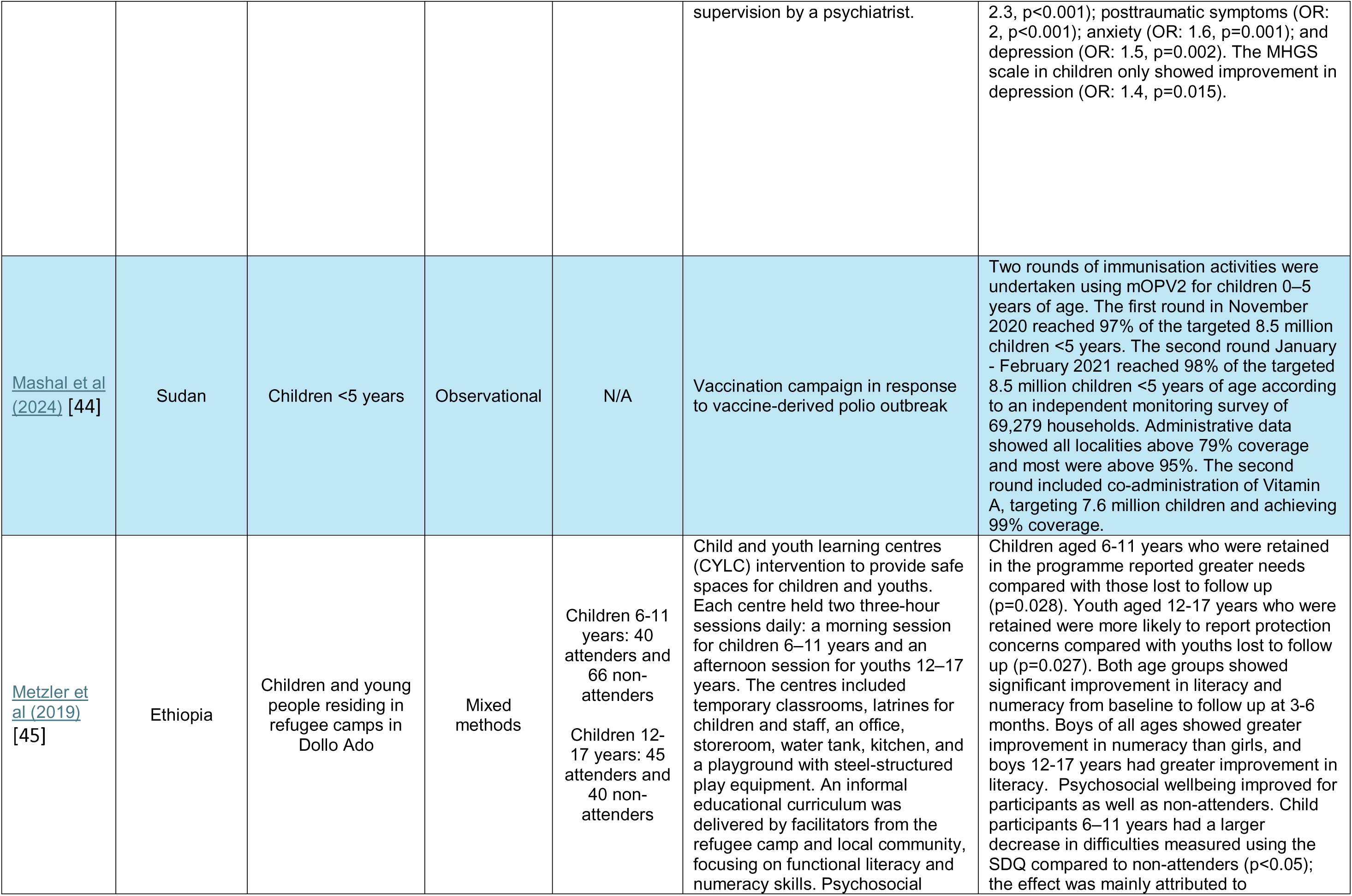
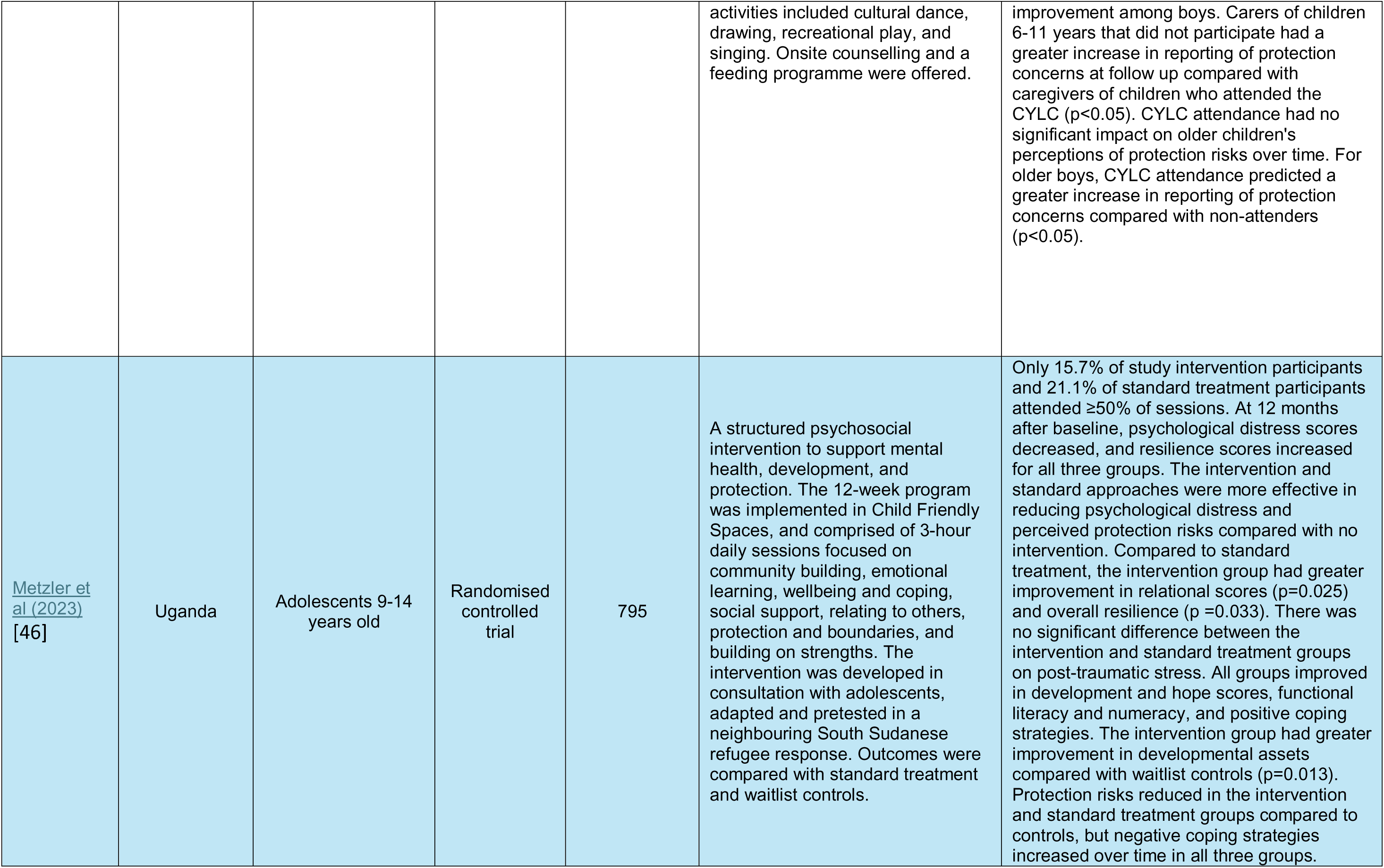
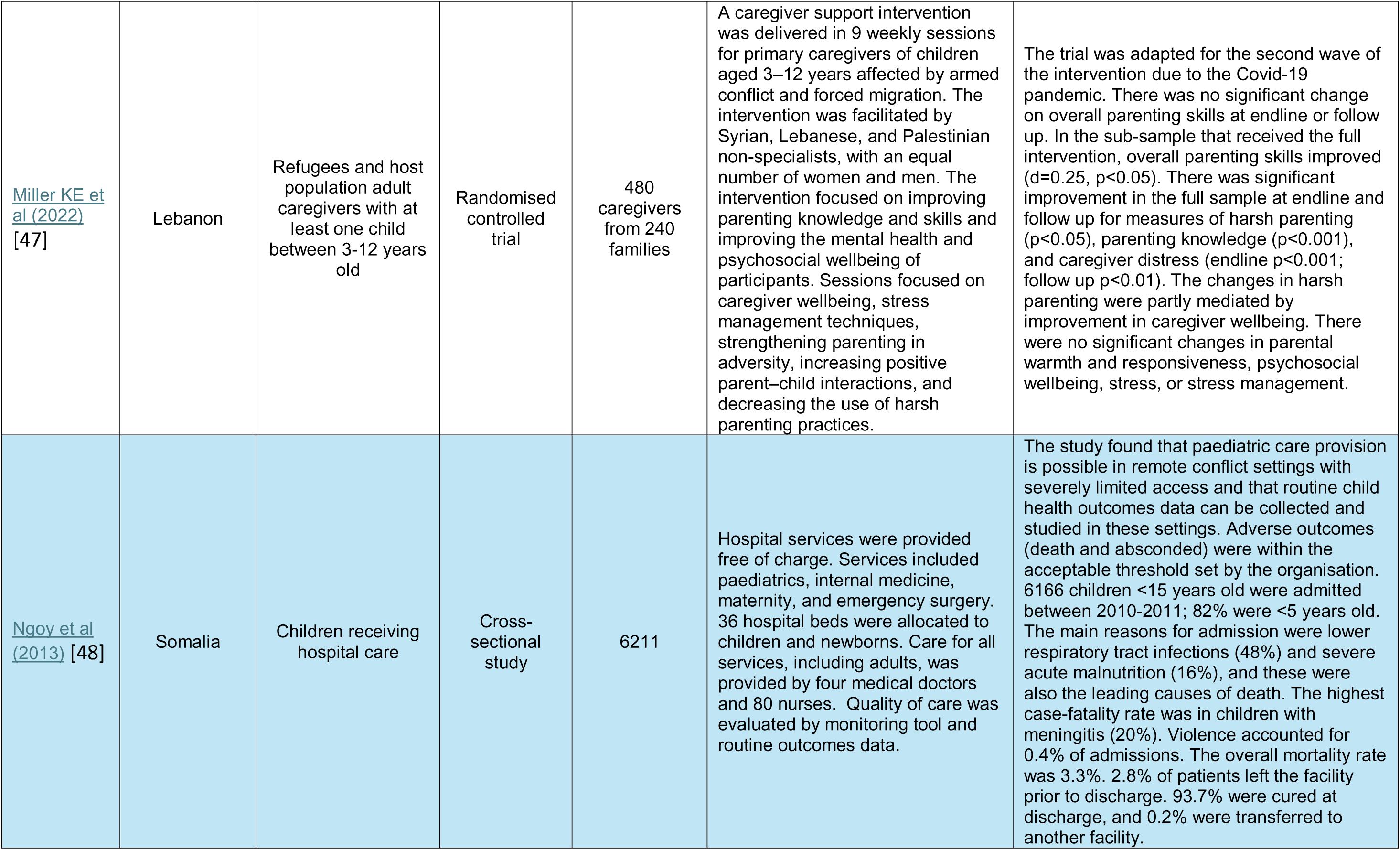
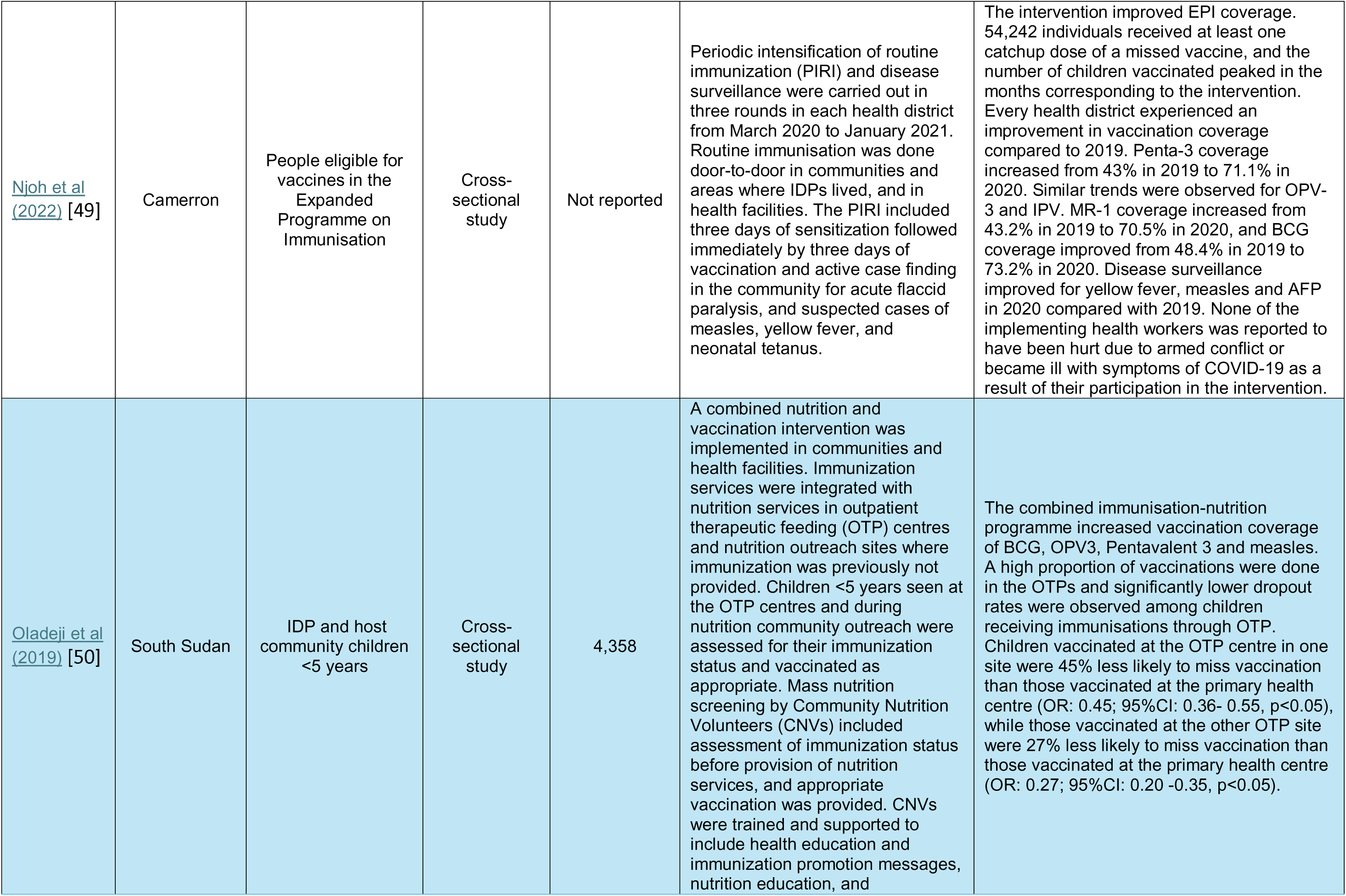
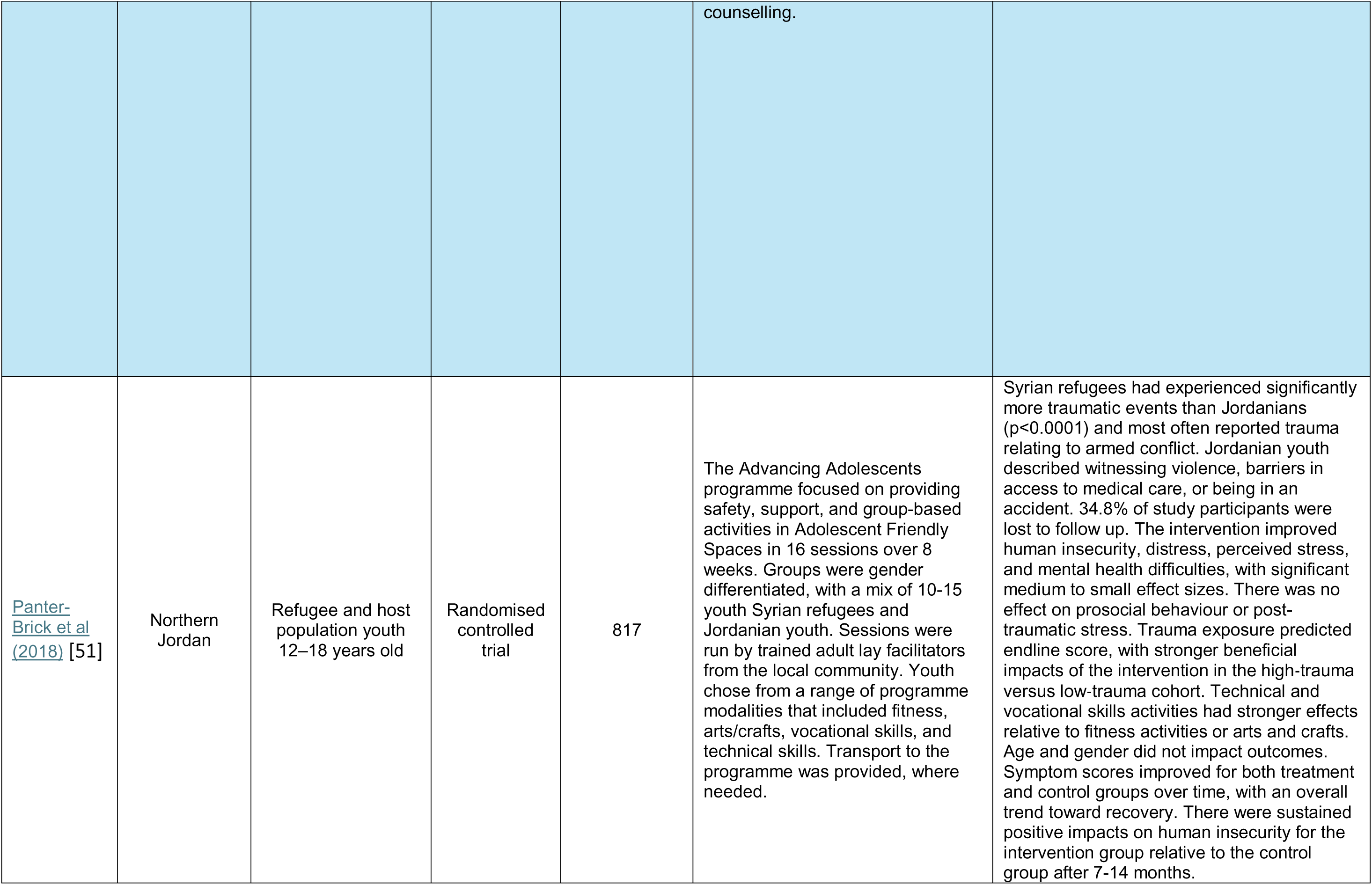
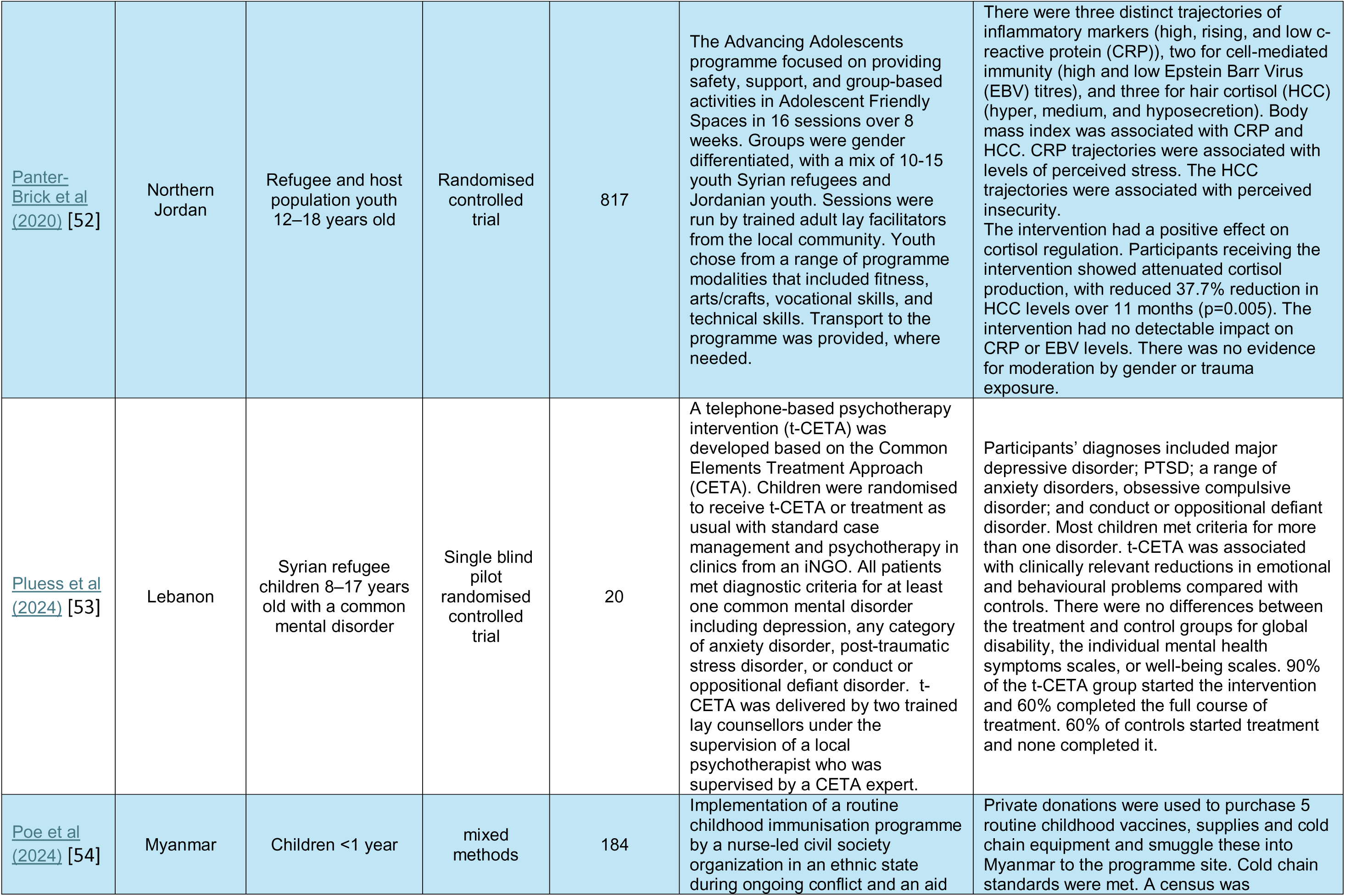
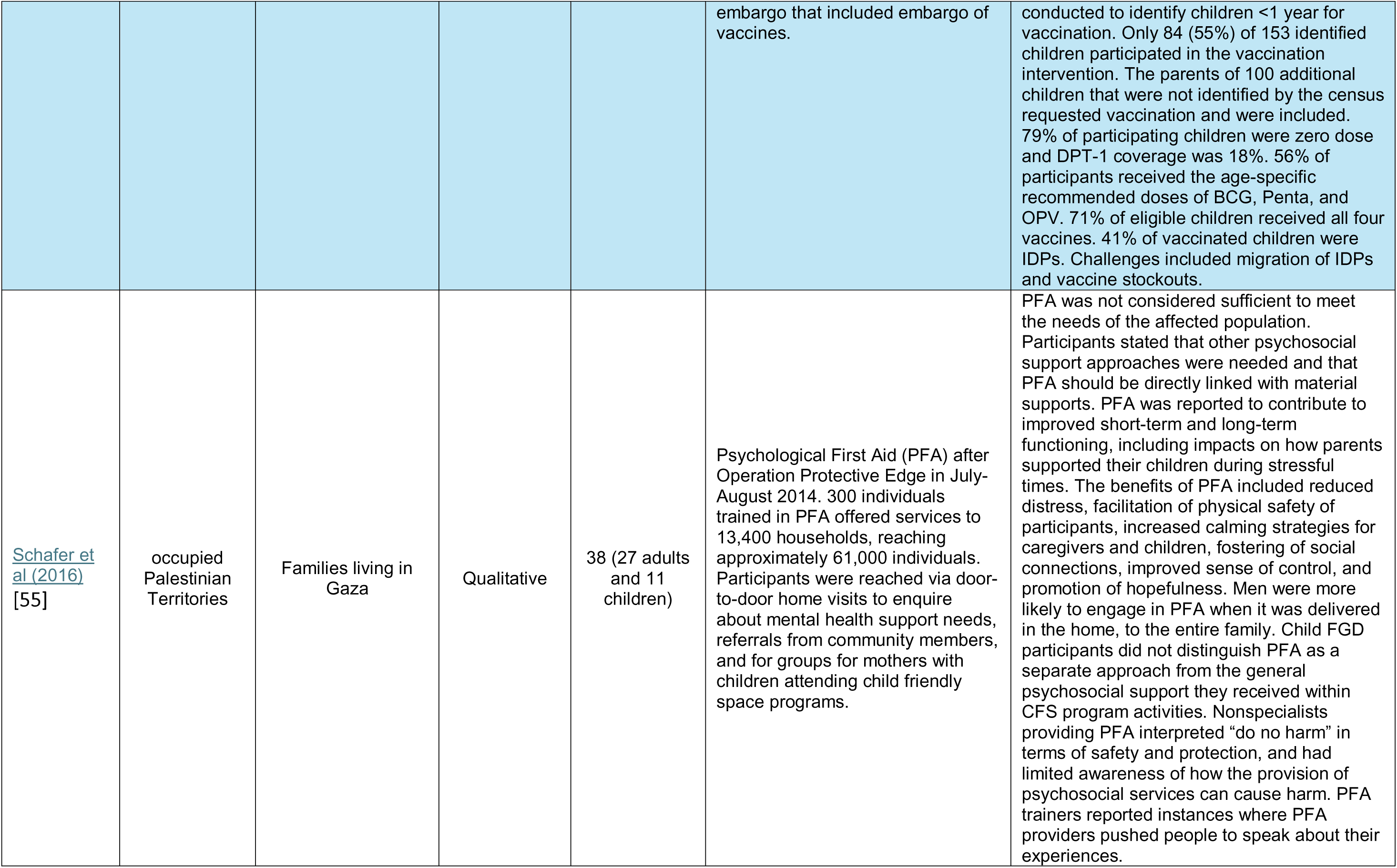
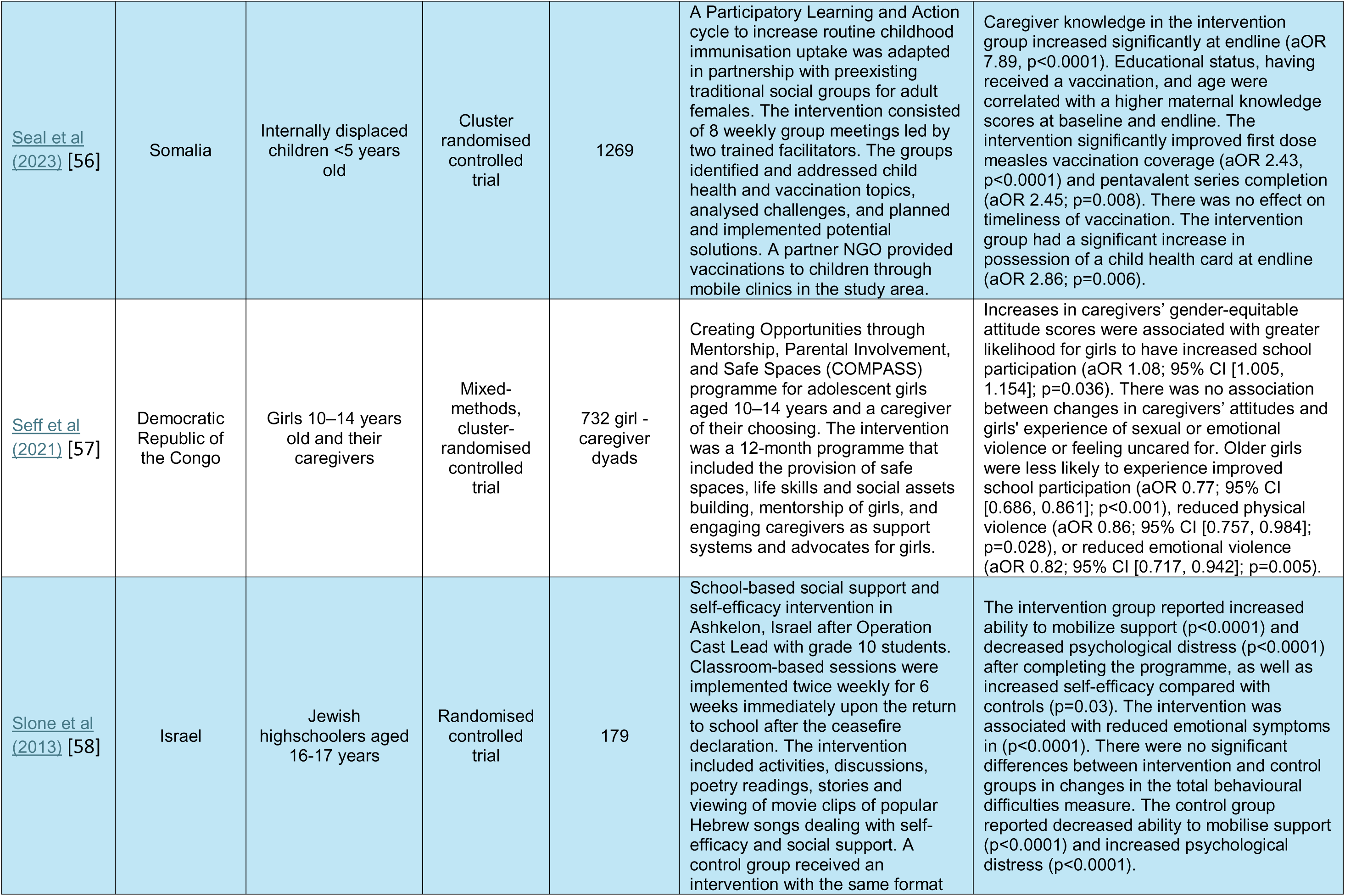
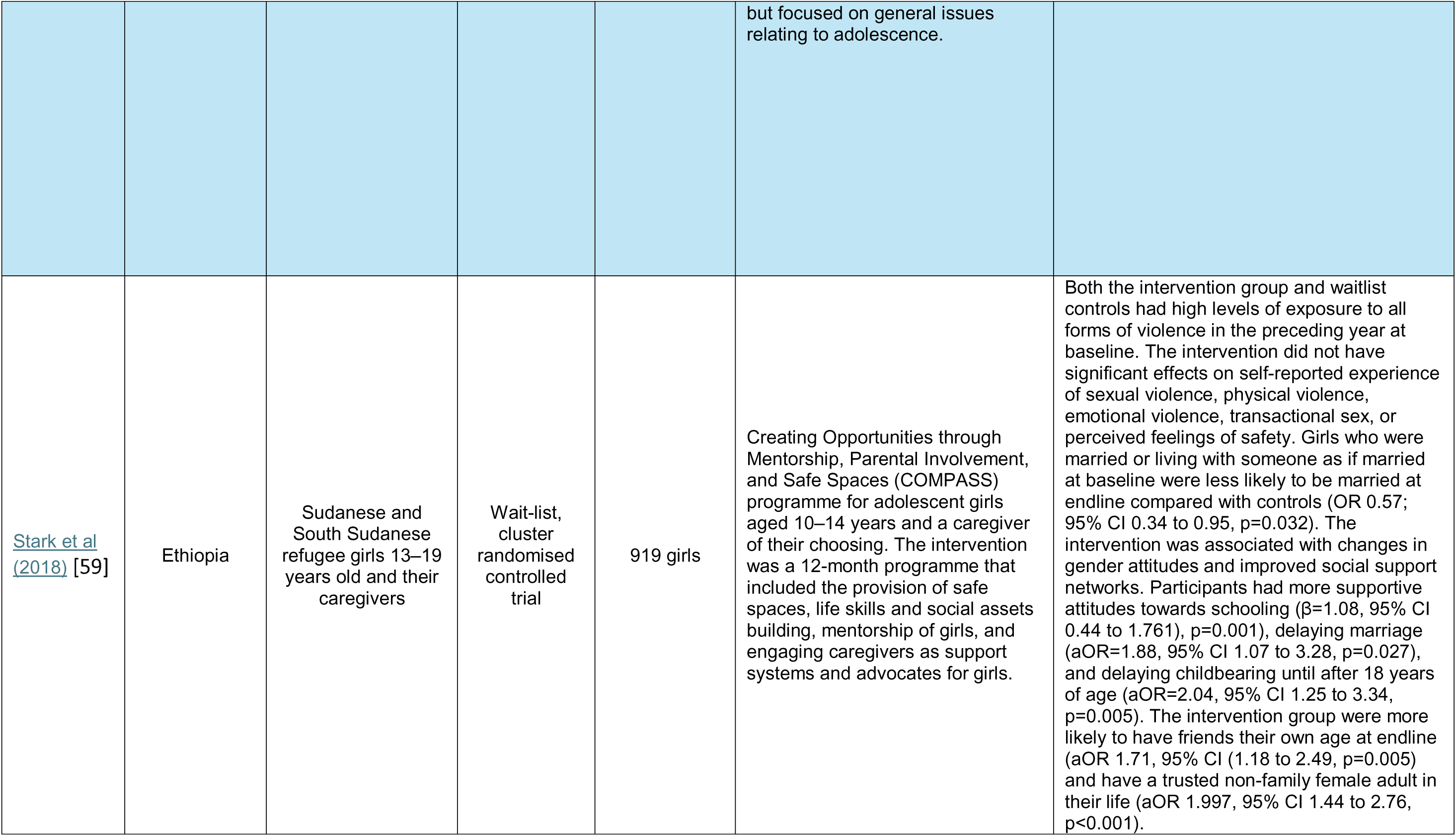
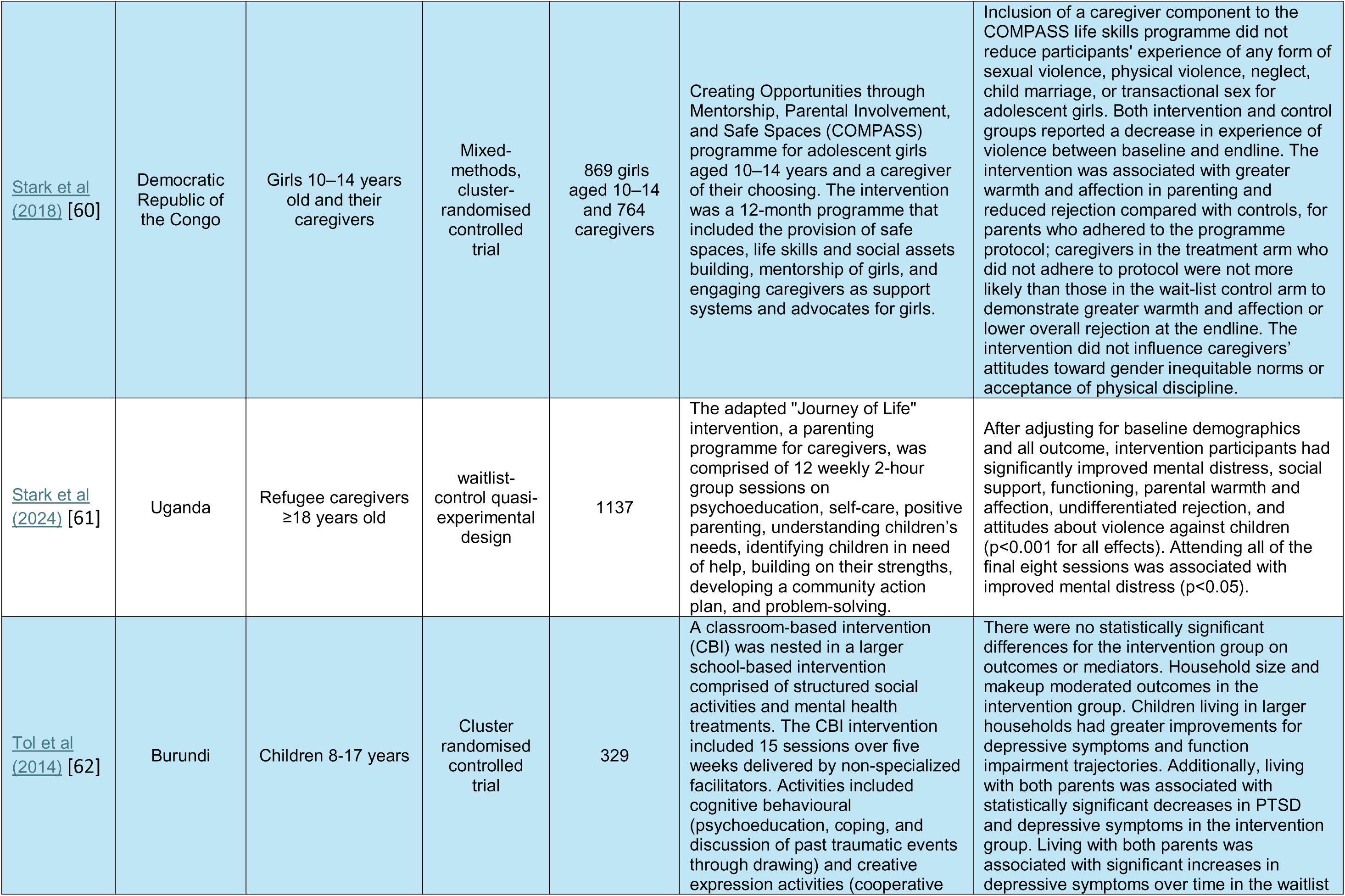
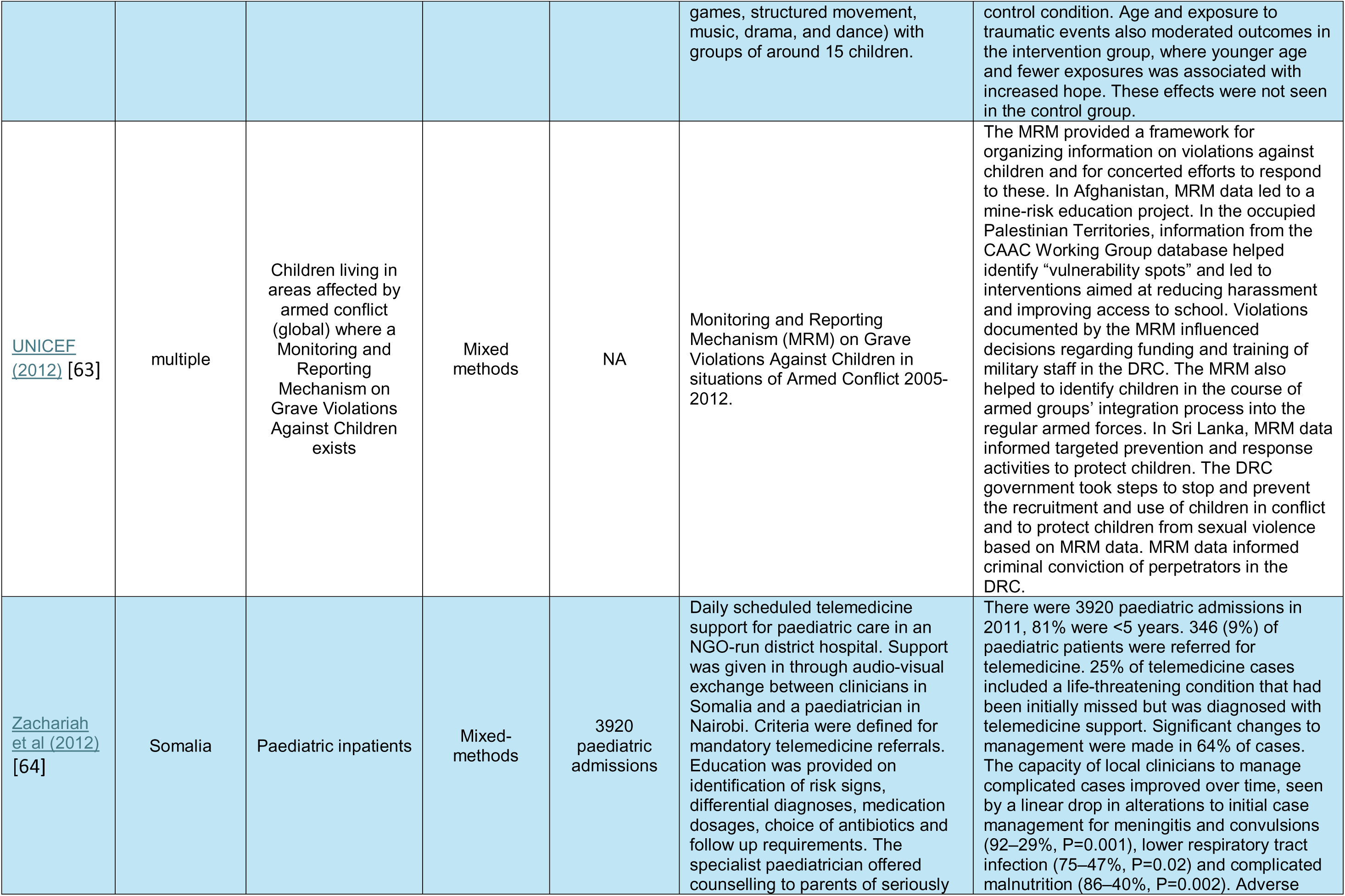
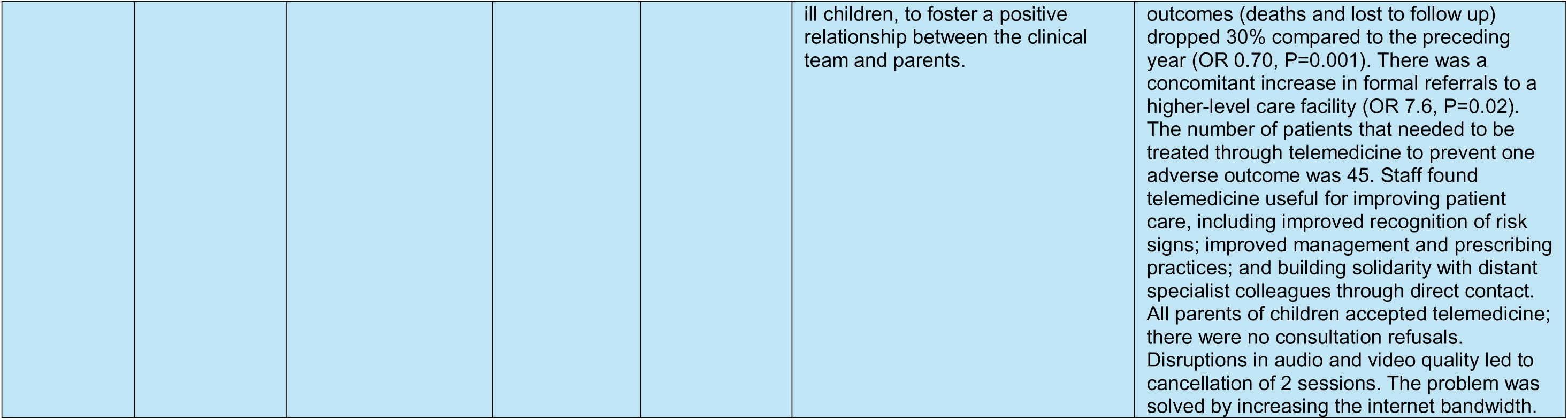
Included studies.

The general characteristics of included studies are presented in Table 2. Twenty interventions were studied in trials, seventeen of which were randomised controlled trials. The trials were almost exclusively focused on mental health and psychosocial support (MHPSS) (N = 16/51) and/or child protection (N = 9/51) and seven included a parenting component. Fourteen trials were conducted with displaced populations. Other study designs included mixed methods (N = 12/51), before and after observational (N = 8/51), cross-sectional (N = 4/51), qualitative (N = 3/51), descriptive (N = 2/51), cohort (N = 1/51) and case-control (N = 1/51). All three qualitative studies were focused on child protection, parenting, and/or MHPSS interventions. All of the studies that included measures of access to services focused on somatic paediatric health interventions.

**Table 2.**
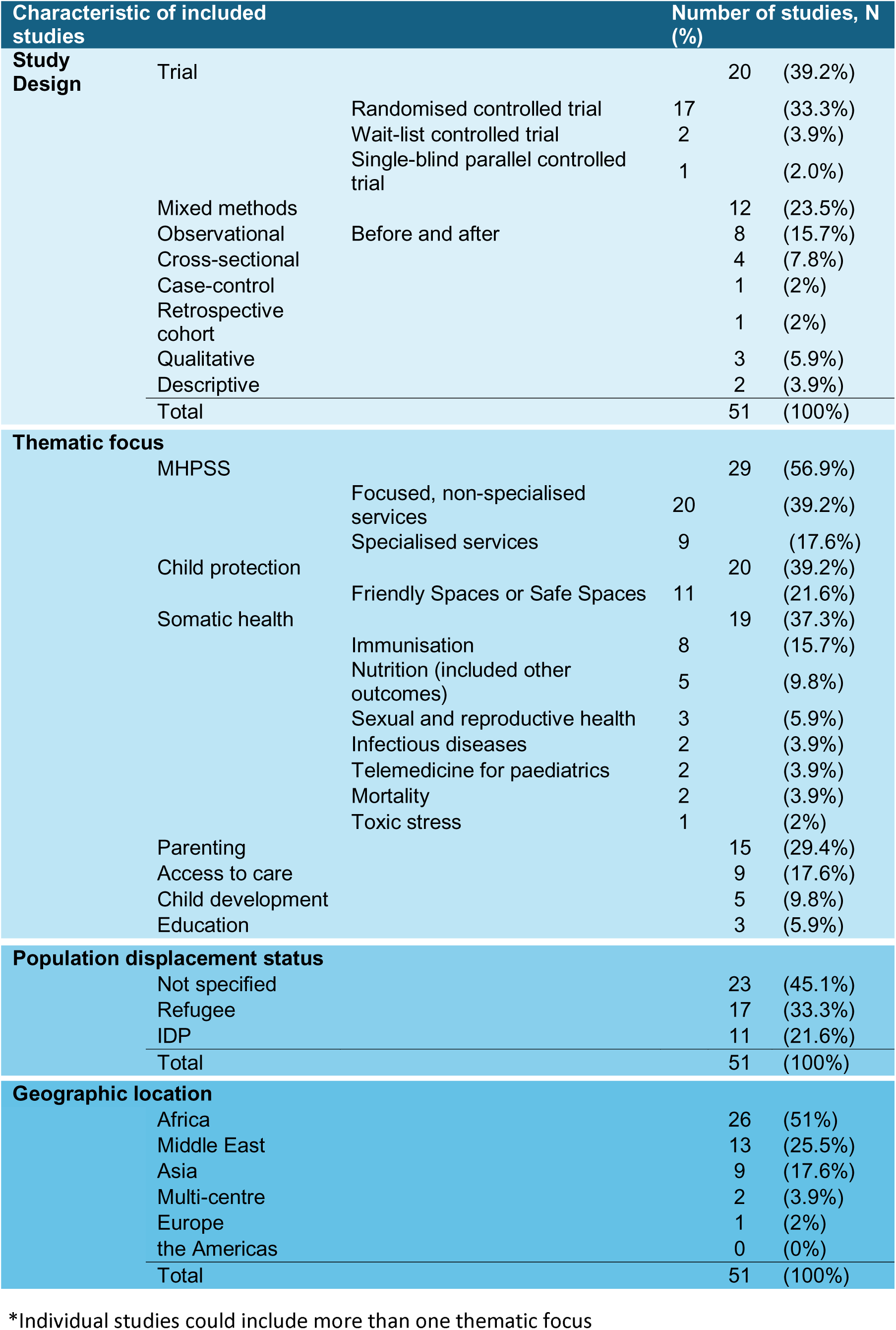
Overview of included studies.

Approximately half of studies were undertaken in Africa (N = 26/51) [14, 22–24, 29, 31, 35, 37–40, 42–46, 48–50, 56, 57, 59–62, 64], followed by the Middle East (N = 13/51) [19, 21, 25, 27, 30, 34, 41, 47, 51–53, 55, 58], Asia (N = 9/51) [15–18, 20, 28, 32, 36, 54], and Europe (N = 1/51) [26]. Two studies were of multi-centre interventions, one with sites in Africa, the Middle East and Asia [33], and the other with a global focus (the Monitoring and Reporting Mechanism (MRM) on Grave Violations Against Children in situations of Armed Conflict) [63]. Figure 2 presents the geographical distribution of included studies. No intervention studies were published in the Americas, despite armed conflicts occurring in ten countries in these regions during the period 2012-2024 [65].

**Figure 2.** **Geographical distribution of included studies** The colour coding applies to entire countries and not regions within countries. All countries that experienced conflict during 2012-2024 are shaded, with darker shading corresponding to the number of studies on child health interventions. This map does not include NATO forces sent from numerous countries to conflicts in other countries. Countries in conflict source: Uppsala Conflict Data Program [65] and the Armed Conflict Location & Event Data Conflict Index 2024 [66].

Studies tended to focus on short-term intervention effects beginning immediately post-intervention, and up 16 months after the intervention was stopped (N = 50/51). The longest follow up period was for a microfinance intervention aimed at improving adolescent MHPSS, development, and protection outcomes in the Democratic Republic of the Congo (DRC) – it was followed up for two years [31].

Over half of the studies focused on displaced populations (N = 28/51): 11 studies amongst internally displaced populations [14, 18, 37–39, 43, 49, 50, 54, 56, 60, 62] and 17 amongst refugees [19, 20, 23, 26, 27, 29, 33, 34, 41, 45–47, 51–53, 59, 61]. Studies of internally displaced people most often included the host population (N=8/11) [14, 38, 39, 43, 49, 50, 54, 62], whereas refugee studies tended to focus on refugees only (N=12/17) [19, 20, 23, 26, 27, 29, 33, 45, 46, 53, 59, 61]. Nearly half (N=8/17) of the refugee studies described interventions in Lebanon and Jordan [19, 27, 34, 41, 47, 51–53].

### Mental health and psychosocial support interventions

Over half of the included studies focused on MHPSS (N=29/51) [15–19, 23–28, 31–34, 36, 41, 43, 45–47, 51–53, 55, 58, 59, 61, 62]. Child protection (N=20/51) [18, 26, 28, 29, 31, 33, 34, 37, 41, 42, 45, 46, 51, 52, 55, 57, 59–61, 63] and parenting support (N=15/51) [19, 23, 25–28, 32, 37, 41, 47, 55–57, 60, 61] were overlapping areas of focus with studies frequently measuring outcomes in two, or all three of MHPSS, child protection, and parenting support. The MHPSS interventions were primarily group-based interventions (N=22/29) with widely varying designs, including: modified written exposure therapy [15], memory training techniques [16, 17], psychoeducation and support for coping and emotional and behavioural regulation [19, 27, 43, 46, 62], remotely delivered psychotherapy [53], Eye Movement Desensitization and Reprocessing-based group therapy [24], narrative therapy [24], social skills building and self-efficacy training [58], Baby Friendly Spaces [23], Child/Adolescent Friendly Spaces or Safe Spaces [18, 33, 34, 45, 51, 52, 59], Psychological First Aid (PFA) [55], reintegration programmes for children associated with armed forces or armed groups [36], mobile gender-based violence (GBV) services [41], a microfinance programme [31], and interventions for caregivers focused on parenting skills and psychoeducation [25–28, 32, 47, 61]. Remarkably, 25 of the 29 interventions reported clear positive mental health and/or psychosocial support outcomes amongst children participating in the intervention. Of these, only one third (N=8/25) of interventions were delivered by staff with mental health expertise [16, 17, 23, 24, 32, 41, 58, 61]; the remaining 17 interventions were delivered by non-specialists [15, 18, 19, 25–28, 31, 33, 45–47, 51–53, 55, 59].

A series of small studies explored rapid, light-touch caregiver support group sessions focused on parenting skills and emotional self-regulation [25, 26, 32]. One programme in Palestine, inspired by an intervention in Syria where informational leaflets were distributed with bread, involved a single 2-hour parenting and emotional self-regulation information session delivered by non-specialists to 120 caregivers [25]. At three month follow up, participating caregivers reported significantly improved parenting and family adjustment scores, as well as improved symptoms of emotional problems, reduced hyperactivity/inattention, improved prosocial behaviour and reduction in the total difficulties scores in their children when compared to the control group. Two studies evaluated a family skills programme with parallel and joint sessions for caregivers and children 8-15 years old, delivered during three weekly sessions lasting 1-2 hours each (five hours total) [26, 32]. The studies evaluated outcomes in Afghanistan (N=72) and Afghan refugee families living in reception centres in Serbia (N=25), respectively. At 6-week follow up, caregivers in both sites reported statistically significant improvements in their children’s Strengths and Difficulties Questionnaire scores and improved parenting and family adjustment scores compared with baseline. The intervention in Afghanistan was delivered by a mix of mental health professionals and non-specialists, and the intervention in Serbia by non-specialists. Whilst the sample sizes were small and the follow up short-term, the interventions were deemed to have shown promise for brief, low cost, non-specialist mental health support for children and caregivers affected by conflict.

Four papers evaluated the impact of an intervention package to reduce girls’ experience of violence and increase caregiver and social support networks to protect girls from violence. The intervention was implemented in the DRC, Ethiopia, and Pakistan [18, 57, 59, 60], and included safe spaces, life skills training, mentorship, and a complementary caregiver programme. Outcomes in the three countries were evaluated by a mixed-methods study (Pakistan) and two randomised controlled trials (Ethiopia and DRC). In Pakistan, the intervention was found to have improved self-esteem, feelings of hope, and social support networks. At endline, participants were also more supportive of girls having the same opportunities as boys, and of girls working outside of the home after marriage [18]. In Ethiopia, participants demonstrated significantly improved social connectedness and school participation; however, there were no reported changes in girls’ experience of sexual, physical or emotional violence [59]. Similarly, in the DRC, the intervention did not demonstrate an effect on reported experience of sexual, physical or emotional violence, neglect, transactional sex, or child marriage; however, significant improvements were observed in school participation, positive attitudes of caregivers towards gender equity, and increase caregiver warmth and affection [60]. Both the intervention group and the waitlist controls in DRC reported significant reductions in experience of physical and sexual violence at endline. In addition, increased gender-equity attitudes of caregivers was positively associated with school participation [57]. This series of studies highlights some nuances in the drivers of violence against children, the role of caregivers, and the role of context on children and young peoples’ risk profiles.

Whilst the majority of interventions targeted the general population of children experiencing conflict, seven studies tested interventions to treat children who had screened positive for post-traumatic stress symptoms or psychiatric distress symptom [15–17, 19, 24, 27, 53]. All seven studies showed significant improvements in psychological symptoms after completion of treatment. The beneficial effects of the therapies were still evident at endline in the six studies that followed up after three months [15–17, 19, 27] and five months [24]. Three of the seven treatment interventions were delivered by mental health professionals or psychosocial support workers with training and experience in the provision of mental health and psychosocial support [15, 17, 24].

### Somatic paediatric and child health interventions

Nineteen (37%) of the 51 studies included a focus on one or more aspects of somatic child health. Nearly half of these were immunisation activities, including periodic intensification of routine immunisation (PIRI) and disease surveillance [49]; an integrated immunisation-nutrition intervention [50]; a mobile health intervention [35]; three vaccination campaigns [21, 39, 44]; a participatory learning and action intervention to increase vaccination uptake [56]; and a grassroots intervention to restore routine childhood immunisation during a humanitarian aid embargo [54]. All of the immunisation activities were found to have improved immunisation coverage. There were no studies on interventions for childhood noncommunicable diseases (NCDs).

The integrated nutrition-immunisation intervention observed lower vaccination dropout and fewer missed vaccinations in children who received immunisations in outpatient therapeutic feeding centres compared with children receiving vaccinations in health centres [50]. A mobile health outreach intervention delivered by local non-governmental organisations (NGO) was successful in vaccinating 51,168 previously unvaccinated children in hard to reach districts in Somalia over a period of 10 months, despite periodic service disruption due to insecurity [35]. The study did not report on vaccination of under-immunised children, or immunisation coverage. Whilst the three vaccination campaigns, the participatory learning and action intervention, the grassroots immunisation activity, and the PIRI intervention led to improvement in vaccination coverage, only one campaign achieved sufficient coverage for herd immunity. The campaign was a government-led, multi-agency response to a vaccine-derived polio (cVDPV2) outbreak in Sudan in 2020 [44].

Adolescent sexual and reproductive health (SRH) interventions (N = 3/51) were focused on access to care, quality of care, child protection and MHPSS. A study in the DRC described contextual and age-specific patterns in experience of sexual violence and access to care at health facilities delivering sexual violence services in two sites, one post-conflict and one experiencing active conflict [42]. In the post-conflict site, patients accessing care were predominantly adolescents and young people (median age 15 years). The study found that sexual violence was more often perpetrated by civilians known to the survivor, patients were more likely to present for care after 72 hours (outside of the timeframe for optimally effective prophylaxis against HIV and sexually transmitted infections), and 38% had associated trauma. The main reason for delayed presentation was fear. In the site with active conflict, facilities received predominantly adult patients and the study outcomes were not disaggregated by age. A study of access to SRH services in Cameroon identified barriers in access to services for young people aged 10-24 years [22]. Young people described barriers to care, including a lack of awareness of available services or how and when to access them, shame, stigma, and fear, insecurity, while health workers cited a lack of expertise in adolescent SRH combined with services that were not designed to serve young people. Adolescent participants in a study of a mobile GBV intervention in Lebanon reported that participation in psychosocial support activities improved their social connectedness, improved their emotional support and regulation, and increased their sense of safety and confidence in public spaces [41].

One study measured alterations in physiological stress responses in relation to an eight-week psychosocial intervention for refugee and host population adolescents [52]. The intervention was implemented in Adolescent Friendly Spaces in Jordan with age-matched and where possible, gender-matched group activities including fitness, arts and crafts, vocational skills, and technical skills. Intervention participants showed significant reduction in hair cortisol production (a marker of neuroendocrine stress response) across the 11-month study period (p = 0.005). The intervention demonstrated no effect on c-reactive protein levels or Epstein Barr Virus antibody titres.

Telemedicine interventions to support colleagues with limited paediatric expertise providing health care to children in settings characterised by insecurity and limited access for humanitarian actors were described in two studies. The interventions used different technologies to connect with colleagues in Syria (Skype and Facebook) [30] and Somalia (Audiosoft) [64]. The study in Somalia evaluated the changes to treatment and associations with patient outcomes compared with the prior year. The intervention, which included daily scheduled support for paediatric services, found that telemedicine support was associated with significant changes to clinical management in 64% of cases (e.g. changes to treatment regimen or medication dosage), and that 25% of cases referred to telemedicine included a life-threatening condition that was initially missed but later diagnosed with telemedicine support. Over time, management of potentially complicated cases was observed to have improved, evidenced by a linear reduction in telemedicine-directed changes to treatment for meningitis and convulsions (p = 0.001), lower respiratory tract infection (p = 0.02) and complicated malnutrition (p = 0.002). The intervention reduced adverse outcomes compared to the previous year (OR 0.70, 95% CI: 0.57–0.88, p = 0.001). The study reported that 45 patients needed to be treated through telemedicine to prevent one adverse outcome (defined as death or loss to follow up). The other telemedicine intervention documented successful provision of paediatric intensive care and nephrology support for a range of clinical decisions, including real-time support for resuscitation using telemedicine. However, the descriptive design did not enable attribution of outcomes to the intervention.

### Child development and disability

One intervention specifically targeted child development. Four studies incorporated one or more measures of intervention impact on child development. A 10-day play-based intervention for Rohingya refugee children aged 5-12 years aimed to support the development of prosocial behaviour and executive functioning [20]. The intervention found improved prosocial behaviours, with the patterns of change differing according to age and place of birth (Myanmar vs refugee camp). The other four studies described child friendly spaces (CFS) in multiple settings [33, 34, 46], and a microfinance programme with adolescents and caregivers in the DRC [31]. The evaluation of the microfinance programme had three study arms: adult caregivers only, integrated adolescent-caregiver, and adolescent only. For all three groups, children showed improved school attendance (p = 0.015) and prosocial behaviour scores (p1=10.032), with the greatest effects observed in the integrated parent-adolescent arm and the adolescent-only arm. The CFS interventions showed variable results. A meta-analysis of observational studies in five countries showed improved developmental assets in CFS attenders in Uganda (d1=10.37, 95% CI 0.15–0.59) and Iraq (d1=10.86, 95% CI 0.18–1.54), which disappeared on pooled analysis of data from the five sites [33]. One of the five sites was not a conflict- or post-conflict setting and accounted for over one-third of the pooled sample (N = 807, 35%). A study of a CFS intervention in Jordan (for children 6-17 years old) observed a decline in CFS attenders’ developmental assets at endline and at 12-month follow up [34]. A CFS intervention in South Sudan (for adolescents 9-14 years old) found that both CFS attenders and waitlist controls had improved developmental assets at 12-month follow up, with CFS attenders showing significantly greater improvement compared with controls (p = 0.013) [46].

Of the 51 included studies, only two addressed an aspect of childhood disability. One intervention measured a mental health-related disability outcome in Syrian refugee children (aged 8-17 years old) receiving a telephone-based mental health intervention compared with treatment as usual with case management and psychotherapy delivered by an NGO [53]. Both groups had reduced disability scores over time, with no difference between the intervention group and the treatment as usual group. The other study was a participatory educational support group programme for the caregivers of children (aged 2-10 years) living with disability in Afghanistan [28]. The intervention consisted of nine sessions aimed at improving caregiver, child, and family well-being. Participants reported significant improvement in quality of life and family functioning at endline across all domains of the Paediatric Quality of Life, Family Impact Module (p < 0.0001). Caregivers reported changes that they attributed to the intervention, including moving from feeling frustration with their child to a feeling value and love for them. Parenting practices shifted correspondingly, with reports of increased patience, kindness and nurturing. Caregivers reported improved emotional self-regulation and described increased inclusive behaviours, including how they called the child (“mighty” vs “disabled”). Some caregivers reported that their children responded with increased participation in family and social life, improved mood, reduced social isolation, and improved mobility, communication, and self-care abilities after participating in the programme.

### Assessment of harm due to interventions

Only 10 of the 51 studies mentioned any assessment of whether the intervention caused harm to participants. In other words, 80% of published studies did not report whether any attempt was made to identify harm to children or families resulting from or related to their participation in the intervention. Only two studies reported adverse events. One was a vaccination campaign, which identified two cases of fever after vaccination; the patients were evaluated, managed symptomatically, and followed up [21]. The other study - of a PFA intervention - reported that some PFA providers had pressured participants to speak about difficult experiences and then failed to address the emotions and thoughts the participant had shared [55]. None of the remaining nine studies that assessed harm to participants reported adverse outcomes due to the interventions.

## Discussion

Our review identified 51 intervention studies, including several thoughtful studies that sought to address nuanced and upstream determinants of child health such as the role of caregivers and family, child and adolescent development, children’s agency, and economic assets. Whilst the literature on child public health in conflict settings remains sparse, there is a notable increase in both the number of intervention studies published in recent years as well as the use of more rigorous methods to evaluate interventions in conflict settings, particularly for MHPSS interventions. This is an improvement compared to the findings of a large, general systematic review of the effects of armed conflict on child health and development from 1945-2017, which identified mostly cross-sectional or descriptive studies [2].

Our review identified a range of promising MHPSS, parenting and child protection interventions, two-thirds of which were delivered by non-specialists. While nearly half of somatic paediatric interventions were immunisation activities, reports also described interventions for adolescent sexual and reproductive health, toxic stress, and telemedicine interventions to improve access to and quality of paediatric health services. The geographical distribution of interventions is similar to previous studies, with notable gaps in evidence for the Americas, North Africa, West Africa, South Asia, and Southeast Asia and the Pacific [2, 12]. The geographical pattern of publications may reflect political framing of conflict, especially in Latin America [67]. Research focusing on the health impacts of armed conflict is relatively new, and prioritisation of funding for research in certain contexts is likely to influence the distribution of studies [2].

### First, do no harm

It is concerning that only 10 of the 51 included studies mentioned any assessment of whether the intervention caused harm to participants. Among the 20 interventions that included a child protection component, only two studies mentioned assessment of harm to children from the intervention. These findings suggest that accountability to children in humanitarian response is a low priority; findings which are mirrored by the relative lack of mechanisms to enable accountability to children in the larger humanitarian health architecture [68]. The concern for harm to children caused by humanitarian interventions that were intended to support them is well founded. The risk is long recognised, and standards and guidance have been developed to prevent harm by humanitarian actors, with heavy focus on preventing exploitation and abuse [69, 70]. There may also be unintended harms related to limitations in paediatric or child public health skills and knowledge, limited child-specific resources, and/or lack of adequate support and supervision. An example of the latter was seen in a study included in this review that identified instances where PFA providers encouraged people to speak about their traumas and then failed to support them afterward. A range of challenges relating to quality and safety of services for children have been described elsewhere [71].

Concerns about poor quality of care and barriers in access to health services for children in crisis contexts has led to a call for an intentional, informed and collaborative effort to improve child health services, beginning with a recognition of Humanitarian Paediatrics as a field of specialty within public health [71]. Two of the studies included in this review focused on telemedicine interventions, which aimed to do exactly that: improve the quality of paediatric care in settings with limited child health expertise, limited resources and lack of access due to insecurity. Whilst it does not address the underlying issues of resource distribution, telemedicine can be seen as a kind of palliative measure to improve existing services. Telemedicine carries a particular set of risks to patients that relate to patient confidentiality, the use of digital technology to collect and share sensitive personal information, and the potential to reinforce colonial power and resource differentials [72, 73]. It is noteworthy that neither of the reports of telemedicine interventions mention assessment of risk, mitigation of risk, or occurrence of harms to children who received telemedicine support. One of the studies described using Skype and Facebook Messenger for communication with colleagues; the study took place before the General Data Protection Regulation (GDPR) came into effect, however it is important to note that this practice violates the GDPR [74]. Even carefully maintained and secured databases are vulnerable, as demonstrated by the 2022 cyber-attack on an International Committee of the Red Cross (ICRC) protection database [75].

### Improved evidence and persisting evidence gaps

Over half of included studies and 95% of trials focused on MHPSS, child protection and/or parenting interventions, suggesting a growing demand for evidence to inform interventions in these sectors. A range of MHPSS, child protection, and parenting interventions were successfully delivered to conflict-affected populations, some of which were living in insecure environments due to conflict. Non-specialists (e.g. doctors, clinical officers, nurses, midwives, or community health volunteers) delivered a large proportion of these interventions with supervision and support from mental health specialists, and most of these interventions showed promising results. These findings lend support to the current approach of larger foundational non-specialist interventions for basic needs, self-efficacy, and strengthening of community support, with respectively smaller scale non-specialised and specialised services [76]. The predominance of non-specialised MHPSS interventions is also consistent with a recent review of humanitarian health interventions that was not child-specific [77], however our review findings differed with respect to intersectoral interventions. We found that interventions for children and adolescents frequently took an intersectoral approach, combining MHPSS with elements of child protection and/or parenting support. However, this may simply reflect the humanitarian architecture, as child protection actors often take a leading role in MHPSS interventions [78].

Our review findings demonstrate that there is a long way to go to meaningfully incorporate the full breadth of child public health into humanitarian responses for children. Although studies of interventions focused solely on nutrition were excluded due to existing reviews on the topic, we included five studies reporting nutrition outcomes that were measured alongside other measures of child public health. The integration of nutrition with other study topics indicates a recognition of the importance of nutrition on child health. Additionally, the large proportion of interventions focused on parenting indicates a recognition of the role of caregivers and other adults in influencing child health and protection outcomes. Whilst this is encouraging, our review identified a lack of studies on childhood NCDs, child development, and disability interventions. The lack of studies on childhood NCDs is consistent with a recent systematic review on NCD interventions for women and children [79], and mirrors the lack of routine data collection on childhood NCDs by humanitarian actors in conflict settings [5]. Further, Munyuzangabo et al identified a lack of evidence for newborn health interventions [12]. Together, these findings indicate a lack of paediatric and child public health expertise among humanitarian actors.

Our review also identified a series of therapeutic MHPSS interventions for children and adolescents with recognised psychological distress or mental health disorders, which showed promising outcomes in the short-term. These therapeutic intervention studies indicate a growing prioritisation of mental health services during humanitarian response; this is an encouraging development, in light of the potentially life-changing and life-long harmful psychological impacts of conflict on children [4]. The frequent overlap of MHPSS, child protection, and parenting also indicates an increased focus on the social determinants of child health. With the existing funding and access challenges in conflict settings, it is unsurprising that most interventions were short-term or nested within interventions such as safe spaces. There is a need to identify solutions for continuity of therapeutic MHPSS services that are safe, effective, feasible, and sustainable in the long-term. Whilst the success of non-specialist interventions lends support for the use of these interventions in conflict settings, non-specialist services are unlikely to be sufficient for all patients, and risks of harm were identified that should be prevented or mitigated.

Several studies reported on interventions in baby-, child-, and adolescent-friendly spaces and safe spaces. The variable findings of these studies is consistent with other studies on the effectiveness of CFS [80]. Safe and friendly spaces are a mainstay of humanitarian response for children and provide an opportunity to support children’s wellbeing, identify health and protection needs, and link them with relevant services. Studies and reviews on CFS and safe spaces underscore the fundamental role of context in influencing outcomes, and the importance of safety and quality of services to effectively support and protect children [80]. The lack of evidence on long-term benefits does not negate the value of friendly and safe spaces for children and young people. From a child public health approach, even short-term benefits to wellbeing and functional literacy may serve to mitigate the toxic stress impacts of severe adversity on children’s bodies and brains. The study by Panter-Brick et al demonstrates the potential to mitigate toxic stress by an MHPSS intervention in a Child Friendly Space [52].

### Development and disability

Five of the included studies explored child and adolescent development using a mental health approach. No somatic studies explored child development either as an outcome or predictor of child physical health. Child and adolescent development is a major gap in humanitarian health response [81], and the lack of somatic paediatric or child health framing reflects the previously-described lack of both paediatric and child public health expertise amongst actors working in humanitarian settings [71]. This gap in expertise was also demonstrated in a recent scoping review of child public health indicators in fragile, conflict-affected and vulnerable settings [5]. The review did not identify a single child development indicator that is routinely measured by five large operational NGOs; recommended by technical agencies and partnerships; or required by donor agencies [5]. Our initial search identified one study describing three case studies on the implementation of Nurturing Care in conflict-affected children [82], however the study was excluded because it did not report data on children’s or caregivers’ outcomes. Nurturing Care is an approach to mitigate toxic stress and promote healthy child development through focus on good health, adequate nutrition, safety and security, responsive caregiving and providing opportunities for early learning [83]. A study of a CFS intervention demonstrated that harmful physiological responses to conflict-related stressors in adolescents can be treated effectively [52]. Toxic stress is well-described in the paediatric literature and has important implications for children and young people’s physiological, neurological, and social development in the immediate-, short-, medium- and long-term [84]. Whilst the evidence on child and adolescent development in conflict settings remains limited, the studies included in this review make some progress towards documenting this foundational aspect of child health and wellbeing.

Childhood disability remains largely out of focus, with only one study of an intervention to support children with disability [28], and another study which included a measurement of disability related to mental health [53]. There were no interventions on injury care or rehabilitation. The limited focus on childhood disability is also seen in data collection by humanitarian public health actors [5]. The lack of data on disabilities amongst conflict-affected children is deeply concerning. Disabling injuries and illnesses in conflict-affected children are well documented [2]. These impacts on children are not historical. From January – November 2024, an estimated 21,000 children in Gaza sustained war-related injuries, and one-fourth (N = 5,230) of these children require rehabilitation [85]. This equates to an average of 15 children in Gaza sustaining potentially disabling injuries every day – the Protection Cluster in the Occupied Palestinian Territory states that these numbers are thought to be underestimates [85].

Responding safely and effectively to potentially disabling injuries in conflict-affected children requires technical expertise in paediatric trauma, medical equipment that is suitable for children, and understanding of the context-specific determinants of health that may affect the child’s healing and prognosis during care and after discharge [71]. There is an urgent need to improve our understanding of childhood disability in conflict settings and of interventions to support affected children in the short- and long-term. Similar to interventions on child and adolescent development, this will require reliable data. While the availability and quality of child public health data are limited, routine situation and outcome data on child public health are collected in most settings by humanitarian actors, demonstrating that it is feasible to collect routine data in most contexts despite of contextual and funding challenges [5].

The intervention by Evans et al [28] included in this review is an excellent example of using a child public health approach to protect and promote the health of children with disabilities. The intervention was designed in recognition that child health, development and safety are influenced by social dynamics within the home and in the community, perceived value of the child, caregiving practices, and caregiver mental health. Participants reported extraordinary improvements in the abilities of their children after the intervention. For example one child that was previously immobile began to move independently, and another child who was nonverbal began to speak and also play with other children. Such profound improvements in children’s health after positive changes in the social environment and with loving caregiving are a familiar observation for paediatricians and are well-described in the paediatric literature, most notably among children adopted from orphanages [86]. There is a great, yet untapped potential to meaningfully support child health and development for all children in conflict-affected settings, including the most severely impacted children, through the use of child public health.

## Limitations

The findings of this review are likely to be limited by reporting bias, including reduced reporting of negative and/or nonsignificant outcomes [87]. Further, many of the studies included in our review measured outcomes based on participant and/or caregiver self-report and are thus prone to response bias. In humanitarian settings, affected populations may feel pressured to participate in studies in order to receive aid, presenting both ethical challenges to research as well as a risk of both selection bias and response bias that may affect study findings. A number of studies included in the review were observational, and several did not include a baseline comparison, limiting the conclusions that can be drawn. Reporting of gender-disaggregated outcomes was variable and thus limited our ability to report on gender-specific outcomes. Finally, the geographical distribution of studies was characterised by a notable lack of studies both within and across regions, especially from the Americas, and in particular Central and South America, as well as West Africa, North Africa, South Asia, Southeast Asia and the Pacific. The geographical gaps limit the visibility of conflict-affected children in the missing areas, and the evidence for interventions to support children may not be generalisable to these contexts. Nevertheless, our review compiles the available recent evidence for child public health interventions in conflict-affected children and may serve as a resource for actors wishing to design and deliver clinical or public health interventions for children.

The review included studies published in the peer-reviewed literature and grey literature. This design likely only captures a fraction of the evidence on the effectiveness and/or impact of interventions that were delivered for children affected by armed conflict during the study period, as many intervention evaluations by implementing and donor organisations are not shared publicly. Whilst our review had no language restrictions, we did not search region-specific databases, which may have limited the number of retrieved records and influenced the geographical distribution of identified studies. The search update did not include searches of the Global Health database or the grey literature, which may also have limited the number of retrieved studies. Lastly, the quality of studies and the certainty of evidence were not assessed, thus limiting the conclusions that can be drawn about the design and impact of the interventions. However, refraining from assessing quality and the certainty of evidence served to reduce bias towards better-resourced studies in more stable settings and improve the external validity of our review.

## Conclusion

The evidence for child public health interventions to support conflict-affected populations remains limited, but the numbers of studies and rigour of study design is improving. MHPSS, child protection, and parenting interventions show promise, and the increasing use of intersectoral approaches for these three thematic areas is encouraging. There are few studies of interventions to support somatic child health, particularly child and adolescent development, NCDs of childhood, and childhood disabilities.

Geographical disparities in evidence persist within regions and globally. Few studies mention whether any assessment was made of harm to participants either caused by or related to the interventions. Given the challenges in delivering humanitarian responses for children, and the well-recognised risk of harm due to humanitarian aid, the lack of focus on safety is deeply concerning. Whilst the majority of intervention studies show promising results, the lack of data about safety suggests that conclusions about effectiveness should be treated with caution. There is an urgent need for evidence on the safety of interventions, medium- and long-term impacts, and interventions to support child and adolescent development, children with disability, and childhood NCDs. Research should be prioritised in geographical areas where data are lacking, and reporting should be age- and gender-disaggregated. The publication of nonsignificant and/or negative outcomes should be encouraged, as these findings provide valuable insights for humanitarian actors delivering interventions for children and families. Finally, Humanitarian Paediatrics should be established as a profession within public health to support safe and effective child public health interventions in conflict-affected populations.

## Supporting information

Figure 1. PRISMA flow diagram

Figure 2. Geographical distribution of included studies

Study protocol

PRISMA_2020_checklist

Analytic codes

Template data extraction form

## Data Availability

All data are provided within the manuscript or supplementary information files.

https://www.crd.york.ac.uk/PROSPERO/view/CRD42022356007

