## Supplementary material for "Child public health interventions for conflict-affected populations: A systematic review": Figure 1. PRISMA flow diagram

**Identification of studies via databases and registers**

**Identification of studies via other methods**

Initial search records identified from:

Websites (n=301)

DuckDuckGo and ReliefWeb (n=200)

Citation searching (n=4)

Search update

Citation searching (n=2)

Records removed before screening:

Duplicate records removed

Initial search (n=962)

Search update (n=24)

Records marked as ineligible by automation tools (n =0)

Records removed for other reasons (n =0)

Initial search: Records identified from:

Databases (n=2467)

EMBASE (n=1216)

Global Health (n=363)

MEDLINE (n=888)

Search update (n=627)

EMBASE (n=71)

MEDLINE (n=556)

**Identification**

Records excluded

Initial search (n=1293)

Search update (n=580)

Records screened

Initial search (n=1505)

Search update (n=603)

Reports not retrieved

Initial search (n=1)

Search update (n=0)

Reports sought for retrieval

Initial search (n=212)

Search update (n=23)

Reports excluded: 429

Reports sought for retrieval

(n=460)

(47 duplicates removed)

Reports not retrieved

(n=0)

**Screening**

Reports excluded:

Initial search (n=98)

38 Wrong outcomes

15 Not a study

13 No age disaggregated data

7 Wrong population

7 Unclear methods

6 Review

5 Wrong age group or adult population

4 Outcomes not associated with conflict

2 Wrong study design

1 Military study

Search update: (n=11)

5 Wrong outcomes

5 No age disaggregated data

1 Conference abstract

Reports assessed for eligibility

Initial search (n=211)

Search update (n=23)

Reports assessed for eligibility

(n=460)

Studies included in initial review (n = 142)

Of these, intervention studies (n=37)

Studies included in database search update of intervention studies only (n=14)

**Total included intervention studies (n=51)**

**Included**
