## Supplementary figures and images for "Child public health interventions for conflict-affected populations: A systematic review"

### Figure 2. Geographical distribution of included studies

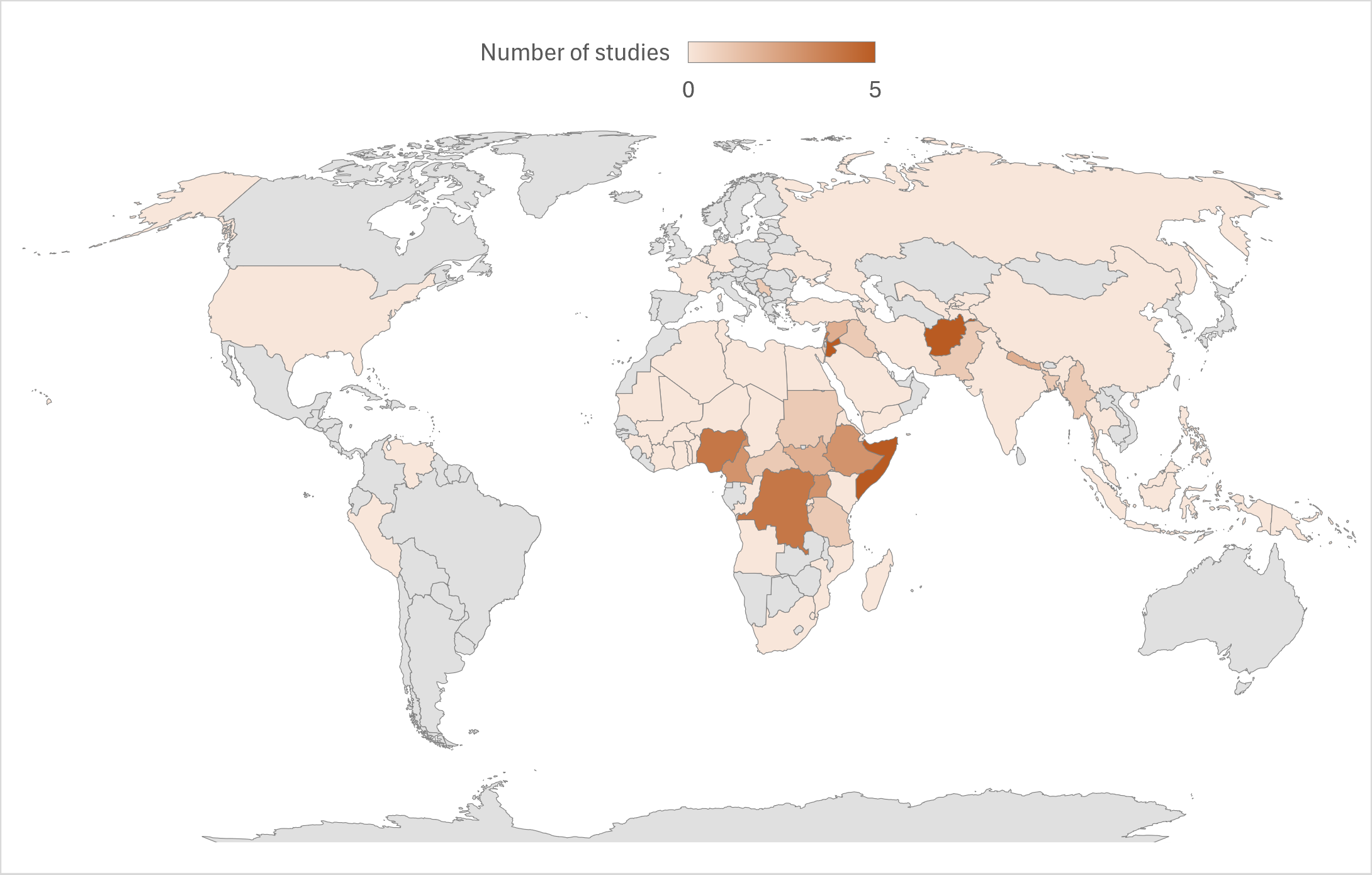
