## Supplementary material for "Child public health interventions for conflict-affected populations: A systematic review": Study protocol

NIHR | National Institute for Health and Care Research

PROSPERO
International prospective register of systematic reviews

### Interventions to protect and promote the health and wellbeing of children and caregivers affected by armed conflict: a systematic review

*Ayesha Kadir, Sneha Krishnan, Catherine McGowan*

**Citation**
Ayesha Kadir, Sneha Krishnan, Catherine McGowan. Interventions to protect and promote the health and wellbeing of children and caregivers affected by armed conflict: a systematic review. PROSPERO 2022 CRD42022356007. Available from https://www.crd.york.ac.uk/PROSPERO/view/CRD42022356007.

#### REVIEW TITLE AND BASIC DETAILS

##### Review title
Interventions to protect and promote the health and wellbeing of children and caregivers affected by armed conflict: a systematic review

##### Review objectives
How do interventions effectively protect and promote health, development, and wellbeing outcomes for children and caregivers impacted by armed conflict and how do they achieve this?
Objectives:
• Identify interventions to protect or promote child health and wellbeing
• Consolidate the evidence on effectiveness of interventions.

##### Keywords
armed conflict, caregivers, child health, conflict, crises, development, displacement, humanitarian, interventions, wellbeing

#### SEARCHING AND SCREENING

##### Searches
**Search terms**
We will use variations of child and armed conflict and setting
(Child* OR infant* OR adoles* OR teen* OR youth OR “young people” OR “young person”)
AND (war* OR conflict* OR violence OR Terroris* OR tortur*)
AND (humanitarian OR cris?s)
**Databases:**

Peer-reviewed lit: MEDLINE, EMBASE, Global Health
Grey literature: ReliefWeb, DuckDuckGo (first 100 results), Global Health, Save the Children, MSF, IRC websites
**Time-period:**
We will include Retrospective and prospective studies
**Publication year**
Studies published between January 1, 2012- September 1, 2022 will be included without any restrictions on language or geography/country.

##### Study design
Inclusion criteria:
Source type
• Original research articles
• Agency technical reports from organisations responding to humanitarian crisis due to conflict
Study design
• Qualitative, quantitative and mixed-method designs
Time period
• Retrospective and prospective
Exclusion criteria:
Source type
• Military studies
• Opinion pieces
• Reviews (systematic and scoping)
• Conference abstracts
• Book chapters
• Case studies on individual or household
• Protocols, methods description only
• Audio/video reports, blog posts
• Social media/media articles
• Guidance documents
Study design
• No research component
• Entirely theoretical

#### ELIGIBILITY CRITERIA

##### Condition or domain being studied
Justification:
Previous reviews have described how armed conflict affects children. This review seeks to provide nuanced understanding of how conflict impacts children and their caregivers and interventions which effectively protect and/or promote child health, development, and wellbeing.
Exposure:
Direct or indirect experience of armed conflict. This includes direct experience of violence from armed conflict as well as being displaced due to conflict, living in an area where armed conflict is occurring or has occurred within the last generation (i.e. the child's parents' generation) or having a first degree relative who has been directly affected by armed conflict.

##### Population
• Children 0-17 years
• Caregivers of children

##### Intervention(s) or exposure(s)
The study will focus on:
• Direct or indirect experience of armed conflict
• Displacement due to armed conflict
• Interventions to promote or protect the health and wellbeing of children affected by armed conflict

##### Comparator(s) or control(s)
No exposure to crises or conflict or violence.

##### Context
• Humanitarian settings due to armed conflict

#### OUTCOMES TO BE ANALYSED

##### Main outcomes
Inclusion studies which report on:
• Morbidity (all cause and disaggregate)
• Disability in childhood
• Child development
• Child mental health
• Caregiver mental health
• Changes in social behaviours and norms in children (e.g. sexual behaviour, child marriage, child labour, etc)
• School attendance and educational attainment
• Mortality
Exclude studies which report on
• Nutrition (multiple reviews exist)
• Maternal, perinatal and neonatal interventions and outcomes (recent review https://PubMed.ncbi.nlm.nih.gov/336082640/ the review found limited literature on neonatal care and postnatal care).

*Measures of effect*
Include studies which provide descriptive and analytical measures of effectiveness of interventions

##### Additional outcomes
None

#### DATA COLLECTION PROCESS

##### Data extraction (selection and coding)
Titles and/or abstracts retrieved using the search strategy will be imported into Covidence and independently screened by two reviewers according to the initial inclusion criteria. If it is unclear

whether an article meets the inclusion criteria it will be carried forward to the next stage. Disagreement will be resolved by discussion or, if not possible, by a third reviewer.
The full text of potentially eligible articles will be retrieved and independently reviewed by two reviewers. In this second round of screening, the full texts of the selected articles will be screened against the inclusion criteria. Disagreement will be resolved by discussion or, if not possible, by a third reviewer.
A data extraction form will be created in Microsoft Excel to extract relevant information including:
• Country, region,
• Type of conflict (internationalised conflict, state-based conflict, non-state armed conflict, one-sided violence, war, as defined in the box above)
• Location of study (refugee or IDP setting, area of active conflict)
• Demographic characteristics of the sample,
• Setting's socio-cultural and gender norms (social-prescribed roles and arrangements, including differences for children, men and women),
• Study design
• Type of interventions, and intervention theory of change
• Causal pathway for interventions and outcomes
• Quotes mentioning effectiveness of interventions or challenges (qualitative only).
• Descriptive and analytical measures of effectiveness of interventions (quantitative only – excluding service attendance data)
• Authors analyses - whether they are from the same organisation that designed the intervention to detect positive biases of reporting results
Any disagreement or discrepancy between the two reviewers will be resolved through discussion with a third reviewer if necessary. Missing data will be solicited from study authors if appropriate.

##### Risk of bias (quality) assessment

Articles will be assessed for quality by two reviewers using the Joanna Briggs Institute checklists for quantitative and qualitative studies based on the study specific design. Discrepancies in scoring will be discussed between the two reviewing authors.

#### PLANNED DATA SYNTHESIS

##### Strategy for data synthesis

Quantitative:
• Quantitative analyses will be conducted using Stata software.
• Basic descriptive statistics will be calculated to summarise information about the study characteristics, samples, and methods.
• Meta-analyses will be performed if relevant
Qualitative:
• Articles will be entered into NVIVO for analysis.
• Thematic synthesis of extracted quotes that mention child or caregiver health needs, interventions, outcomes, contextual factors, and/or causal pathway
• Relevant quotes will be coded using thematic analysis and abstracted into higher order themes.

• An assessment of the quality of findings will be carried out using CERQual (Confidence in the Evidence of Reviews of Qualitative Research).

##### Analysis of subgroups or subsets

• Subgroup analyses will be conducted by age group, sex, and type of conflict

#### REVIEW AFFILIATION, FUNDING AND PEER REVIEW

##### Review team members

**Dr Ayesha Kadir**. Save the Children Fund, UK. England.

No conflict of interest decision selected yet.

**Dr Sneha Krishnan**. OP Jindal Global University. England.

No conflict of interest decision selected yet.

**Dr Catherine McGowan**. LSHTM. England.

No conflict of interest decision selected yet.

##### Review affiliation

OP Jindal Global University

##### Funding source

None

##### Named contact

Sneha Krishnan. F-94 GROUND FLOORJINDAL GLOBAL CITY, SONIPAT, HARYANA, INDIA


#### TIMELINE OF THE REVIEW

##### Review timeline

Start date: 29 August 2022. End date: 31 October 2022

##### Date of first submission to PROSPERO

25 August 2022

##### Date of registration in PROSPERO

26 August 2022

#### CURRENT REVIEW STAGE

##### Publication of review results

The intention is to publish the review once completed.The review will be published in English

##### Stage of the review at this submission

| Review stage | Started | Completed |
| --- | --- | --- |
| Pilot work | | |

| Review stage | Started | Completed |
| --- | --- | --- |
| Formal searching/study identification | | |
| Screening search results against inclusion criteria | | |
| Data extraction or receipt of IP | | |
| Risk of bias/quality assessment | | |
| Data synthesis | | |

##### Review status
The review is currently planned or ongoing.

#### ADDITIONAL INFORMATION

##### PROSPERO version history

- Version 1.0, published 26 Aug 2022

##### Review conflict of interest
None known

##### Country
England, India, Scotland

##### Medical Subject Headings
Armed Conflicts; Caregivers; Child; Humans; Mental Health

##### Details of any existing review of the same topic by the same authors
the review builds upon previous works by the authors:
Kadir A, Shenoda S, Goldhagen J (2019) Effects of armed conflict on child health and development: A systematic review. PLoS ONE 14 (1): e0210071. https://doi.org/10.1371/journal.pone.0210071


##### Disclaimer
The content of this record displays the information provided by the review team. PROSPERO does not peer review registration records or endorse their content.

PROSPERO accepts and posts the information provided in good faith; responsibility for record content rests with the review team. The guarantor for this record has affirmed that the information provided is truthful and that they understand that deliberate provision of inaccurate information may be construed as scientific misconduct.

PROSPERO does not accept any liability for the content provided in this record or for its use. Readers use the information provided in this record at their own risk.

Any enquiries about the record should be referred to the named review contact
