## Supplementary material for "Child public health interventions for conflict-affected populations: A systematic review": Analytic codes

| Physical health | Outcomes reported relating to somatic health, including immunisation, tuberculosis, HIV, other infectious diseases, toxic stress, sexual and reproductive health, child development, disability, mortality, integrated Community Case Management (iCCM), nutrition, and other paediatric patient outcomes |
| --- | --- |
| Mental health | Outcomes reported relating to mental health and/or psychosocial status of children and/or caregivers, including diagnoses of disorders, measurements of psychological distress, functioning relating to mental health, child development, social connectedness, psychosocial support networks, other |
| Social determinants of child health | Caregiver knowledge, parenting practices, child protection, education/schooling, access to care, other |
| Displacement status | Internally displaced (IDP) only, IDP + host population, refugee only, refugee + host population, other, not specified  (by author report) |
| Age group of children targeted | Year or month-range reported |
| Target group gender | Males, Females, Both, Other |
| Study design | Randomised controlled trial, other trial, case-control, cohort, cross-sectional, observational, mixed-methods, qualitative, descriptive, other |
| Mention assessment of harm caused by the intervention | Yes, no |
| Description of harm caused by the intervention | By author report |
| Follow up period | By author report |
| Geographic location of the study | By author report |
